# Engineered antibodies targeted to bacterial surface integrate effector functions with toxin neutralization to provide superior efficacy against bacterial infections

**DOI:** 10.1101/2024.09.23.24313920

**Authors:** Rajan P. Adhikari, Farhang Alem, Daniel Kemboi, Tulasikumari Kanipakala, Shardulendra P. Sherchand, Shweta Kailasan, Bret K. Purcell, Henry S. Heine, Kasi Russell-Lodrigue, Irina Etobayeva, Katie A. Howell, Hong Vu, Sergey Shulenin, Frederick W. Holtsberg, Chad J. Roy, Ramin M. Hakami, Daniel C. Nelson, M. Javad Aman

**Author notes:** equally contributed (Rajan P. Adhikari, Farhang Alem).

## Abstract

Anti-bacterial monoclonal antibody (mAb) therapies either rely on toxin neutralization or opsonophagocytic killing (OPK). Toxin neutralization protects the host from toxin-induced damage, while leaving the organism intact. OPK inducing antibodies clear the bacteria but leave the released toxins unencountered. Infection site targeted anti-toxin antibodies (ISTAbs) that we report here addresses this binary paradigm by combining both functionalities into a single molecule. ISTAbs consist of cell wall targeting (CWT) domains of bacteriophage endolysins fused to toxin neutralizing mAbs (IgG). CWT governs specific binding to the surface of bacteria while the IgG variable domain neutralizes the toxins as they are released. The complex is then cleared by phagocytic cells. As proof of concept, we generated several ISTAb prototypes targeting major toxins from two Gram-positive spore forming pathogens that have a high clinical significance; *Clostridium difficile*, causative agent of the most common hospital-acquired infection, and *Bacillus anthracis*, a Category A select agent pathogen. Both groups of ISTAbs exhibited potent toxin neutralization, binding to their respective bacterial cells, and induction of opsonophagocytosis. In mice infected with *B. anthracis*, ISTAbs exhibit significantly higher efficacy than parental IgG in both pre- and post-challenge models. Furthermore, ISTAbs fully protected against *B. anthracis* infection in a nonhuman primate (NHP) aerosol challenge model. These findings establish that as a platform technology, ISTAbs are broadly applicable for therapeutic intervention against several toxigenic bacterial pathogens.

## Introduction

The success of antibiotics effectively ended the use of convalescent serum therapy for bacterial infections. However, emergence of multiple antibiotic-resistant pathogens, coupled with success of immunotherapeutics in oncology and rheumatology and advancements in biological manufacturing processes, have renewed interest in antibody-based therapeutics to address bacterial infections (*1*). Monoclonal antibodies (mAbs) can act by neutralizing secreted virulence factors such as toxins, or through bactericidal functions or opsonophagocytic killing (OPK) upon binding surface antigens. mAbs offer many unique advantages over conventional antimicrobials by *i)* being highly target-specific, thus sparing the natural microbiome, *ii)* exerting less selective pressure for development of resistance than antibiotics, and *iii)* having superior safety, tolerability, and pharmacokinetic profiles over most antibiotics. Furthermore, their complementary mechanism of action can exhibit synergy with antibiotics (*2*).

As a proof of concept, we chose two clinically significant spore-forming Gram-positive bacteria, *B. anthracis* and *C. difficile*, to generate prototype engineered mAbs referred to as “Infection Site Targeted Anti-toxin Antibodies (ISTAbs)”, to integrate both toxin neutralization and OPK into a single molecule. Toxins are critical for pathogenesis of both bacterial species (*3-6*). Virulent *B. anthracis* encodes two bicomponent protein exotoxin complexes, collectively called “anthrax toxin”, consisting of the protective antigen (PA), which mediates binding to cell surface, along with a catalytic component. The catalytic components of the lethal toxin (LT) is the Zn^2+^ protease lethal factor (LF) (*7, 8*), while the adenylyl cyclase edema factor (EF) serves as the catalytic component of edema toxin (ET), (*9*), which elevates cellular cyclic AMP levels. PA delivers LF and EF to the cytosol and is therefore a key target for immunotherapy (*4-6*). Similarly, the pathogenesis of *C. difficile* is largely driven by GTPase-inactivating toxins TcdB and TcdA (*3*). To-date, three FDA approved mAbs are available for bacterial infections and all are anti-toxin antibodies. Raxibacumab and obiltoxaximab, approved in 2012 and 2016 respectively, both target PA. Bezlotoxumab, approved in 2016, targets *C. difficile* TcdB. At least five mAbs are in human clinical trials that target surface epitopes of *Staphylococcus aureus* or *Pseudomonas aeruginosa*, mediating OPK (*10*). However, to date no bifunctional antibodies have been reported that combine toxin neutralization with OPK activity.

To target anti-toxin mAbs to the bacterial surface, which is required for inducing OPK, we exploited the binding properties of bacteriophage-encoded cell wall hydrolases, known as endolysins. Endolysins attack major peptidoglycan bonds, resulting in cell lysis and release of progeny phage (*11*). Endolysins derived from phages that infect Gram-positive hosts often display a modular structure, with one or more N-terminal catalytic domains fused to a C-terminal cell wall targeting domain (CWT) via an 8-20 residue flexible linker. Significantly, through co-evolution with their bacterial hosts, phage have developed CWTs that bind with high-affinity (nM - pM) to species-specific cell wall teichoic acids or carbohydrates in order to achieve efficient host lysis. Notably, fusion with endolysin CWTs can target heterologous proteins to the surface of target organisms, such as heterologous endolysin catalytic domains (*12-15*), green fluorescence protein (GFP) (*16*), or bacteriocins (*17*). Endolysins from *Bacillus* phage have well-defined CWTs that typically bind the *B. anthracis* neutral polysaccharide composed of galactose, N-acetyl-glucosamine, and N-acetyl-mannosamine (*18-20*). Well-characterized *B. anthracis*-specific endolysins, such as PlyB (*21, 22*), PlyL (*18*), and PlyG (*18, 20, 23-27*), bind to and lyse *B. anthracis in vitro* and *in vivo.* Unlike *B. anthracis* targeting endolysins, the surface epitope for endolysins that target *C. difficile* remains unknown. Nonetheless, of the half dozen studied endolysins that target *C. difficile,* all possess specificity towards *C. difficile* with little to no binding observed in other bacterial species (*28*).

In the present study, the species-specificity of endolysin CWTs is harnessed to drive anti-toxin IgGs to the surface of *B. anthracis* or *C. difficile* bacteria. Engineered chimeras of well-defined and validated toxin-specific mAbs (W2, M18, and AVP-21D9 for *B. anthracis*, and anti-TcdA (6G5) and anti-TcdB (9H10) for *C. difficile*) were generated by fusing the C-termini of their heavy chains(HC) with either *B. anthracis*-specific or *C. difficile*-specific endolysin CWTs (PlyB, PlyG, and PlyL for *B. anthracis*, and 630A and CCL2 for *C. difficile*) (Fig. 1A), creating various ISTAbs (Fig. 1B). As shown in Fig.1C, ISTAbs are expected to bind to the bacterial cell wall, capturing the released toxins and inducing clearance of the bacteria-toxin complex by neutrophils. Candidate ISTAbs were evaluated for the ability to target IgGs directly to the bacterial surface via the CWT domain and benchmarked against parental IgGs to demonstrate retained binding to toxin, toxin neutralization activity, binding to FcγR, and promotion of OPK against *B. anthracis*. The efficacy of lead ISTAbs in comparison to parental IgGs against both pre- and post-*B. anthracis* challenge was evaluated in the mouse model of anthrax. ISTAb efficacy was also evaluated using the non-human primate (NHP) aerosol model of anthrax. Furthermore, for *C. difficile*, the *in vivo* efficacy of lead ISTAb compared to parental IgG was evaluated using the hamster challenge model. In summary, data presented here establish the proof-of-principle for a novel immunotherapeutic concept that should prove applicable to a wide range of toxin-driven bacterial infections.

**Fig. 1:**
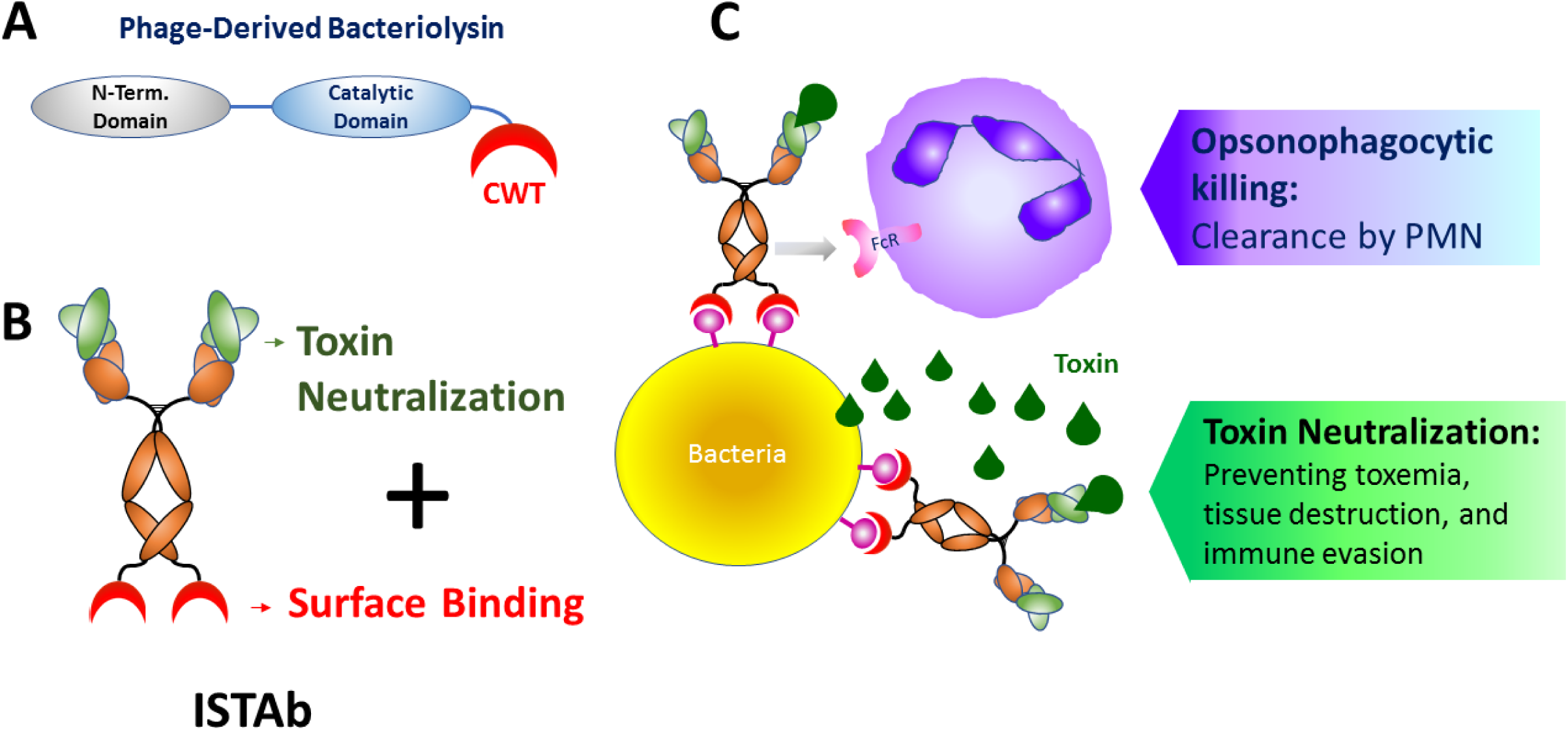
ISTAb Technology. **(A)** Cartoon showing phage-derived bacteriolysin protein with N-terminal domain, catalytic domain and cell wall targeting (CWT) domains. **(B)** Cartoon showing an ISTAb construct. A species-specific cell surface binding region CWT is fused to a monoclonal anti-toxin IgG via a linker sequence. **(C)** Diagrammatic representation of ISTAbs purposed ability to neutralize toxin and prevent toxemia, tissue destruction and immune evasion. Also, its ability to clear bacterial-immune complex via opsonophagocytic killing.

## Results

### Identification of CWTs for ISTAb construction

The γ phage has been used by the CDC for many years as a diagnostic for anthrax because it specifically lyses *B. anthracis* with no activity against non-*B. anthracis* strains, except for *Bacillus cereus* 4342, a *B. anthracis* transition state strain (*20*). Significantly, the γ phage and its endolysin, PlyG, share the same host range. The specificity of PlyG is dictated by its CWT domain, a lectin-like binding domain that recognizes *B. anthracis* (*18-20*). In order to identify *B. anthracis-*specific binding proteins similar to PlyG CWT, we performed a bioinformatic analysis on CWT domains of all endolysins contained in sequenced *B. anthracis* phage and of autolysins of *B. anthracis* strains. The analysis revealed 10 endolysin CWT families containing the archetypical endolysins PlyG, PlyPH, PlyCherry, PlyWBeta, AmiBA2446, PlyL, PlyAP50, PlyTsamsa, PlyB, and PlyBeta. However, several of these endolysins shared high sequence identity (84-99%), narrowing our search down to five unique CWTs with lower sequence identity (range 12-66%) (Fig 2A). All five *B. anthracis* CWTs were recombinantly expressed in *E. coli* and purified. The PlyL- and PlyG-CWTs were expressed with high yields, whereas those of PlyTsamsa and PlyB had very low yields, and PlyAP50 did not fold properly. Thus, PlyL, PlyG along with PlyB were promoted as “finalist” CWTs for further evaluation against a panel of *Bacillus* species. PlyL and PlyG bound to *B. cereus* 4342, a transition state strain, four attenuated (BSL2) *B. anthracis* strains (Ames35, UM23, Sterne, and ΔSterne), and two fully virulent (BSL3) *B. anthracis* strains (Ames and ASC506), but not to other *Bacillus* species including other *B. cereus* species, confirming the high binding specificity of the CWTs for *B. anthracis* (Fig. 2B). None of the *B. anthracis* strains displayed any autofluorescence, validating that any observed fluorescence signal is the result of CWT binding (Fig. 2C).

**Fig. 2:**
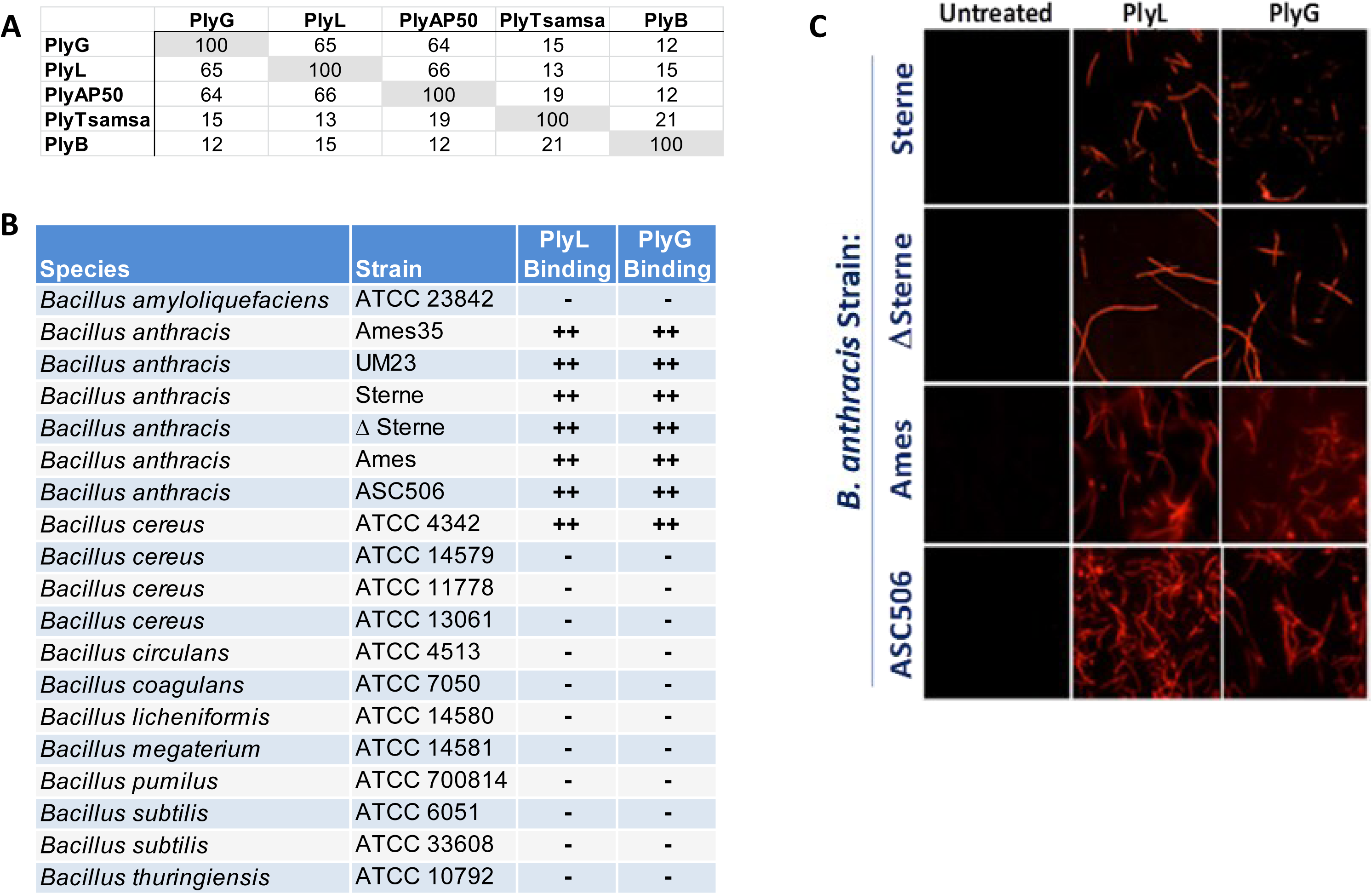
Characterization of endolysin CWTs. **(A)** Sequence homology of five unique *B. anthracis* endolysin CWTs. **(B)** Binding results for PlyL and PlyG CWTs tested against a panel of *B. anthracis* species. **(C)** Fluorescence microscopy image of Alexa Fluor-labeled PlyL and PlyG CWT binding to BSL2 strains (Sterne and ΔSterne) and BSL3 strains (Ames and ASC506) of *B. anthracis*.

### Construction, purification, and characterization of ISTAbs

The mAbs M18, W2, and AVP-21D9 (all IgG1) were selected as the scaffolds for the construction of *B. anthracis* ISTAbs. M18 binds PA with high affinity (*K*_D_ of 35 pM) (*29*), neutralizes the activity of both the lethal toxin *in vivo* (Fischer 344 rat model) and the edema toxin *in vitro* (CHO cells) (*30, 31*), and blocks the binding of PA to fetal Rhesus lung cells (FRL-103) (*9*). W2, a highly neutralizing chimpanzee-derived mAb, binds PA with a *K*_D_ of 40 pM and neutralizes the cytotoxicity of the lethal toxin in a macrophage lysis assay (*32*). AVP-21D9 is a well-characterized human PA neutralizing mAb that provided up to 92% and 67% survival in rabbits and macaques, respectively, against anthrax challenge. Infusion of AVP-21D9 (10 mg/kg) is well tolerated in healthy adults (*33*).

The HC of each antibody (M18, W2, and AVP-21D9) was fused with a *B. anthracis* specific CWT (PlyB, PlyG, or PlyL) and co-expressed with the respective light chain to generate nine different ISTAbs along with their parental IgG. As shown in Fig. 3A, the HC of each ISTAb showed the anticipated larger molecular weight band compared to the parental IgG and exhibited no notable degradation. Using differential scanning fluorimetry (DSF), we evaluated whether the CWTs altered the thermal stability of ISTAbs. Towards this end, we calculated the average melting temperatures (T_m_) using a thermocycler-based method wherein the SYPRO-orange dye binds preferentially to hydrophobic patches that are exposed as the proteins unfold with increasing temperature (Fig. 3B, Supplemental Table S1). M18 IgG and M18-PlyB ISTAb exhibited a T_m_ of 68.5±0.5℃ and 55.5±0℃, respectively, corresponding to a thermal shift of 13℃, while addition of the PlyL or PlyG CWTs to M18 minimally reduced the Tm (<2℃). Similarly, W2-PlyB showed a modest reduction in thermal stability (2.7℃). Addition of PlyL or PlyG-CWTs did not have any significant effect on W2 (Supplemental Table S1) or AVP-21D9 stability (Fig. 3B).

**Fig. 3:**
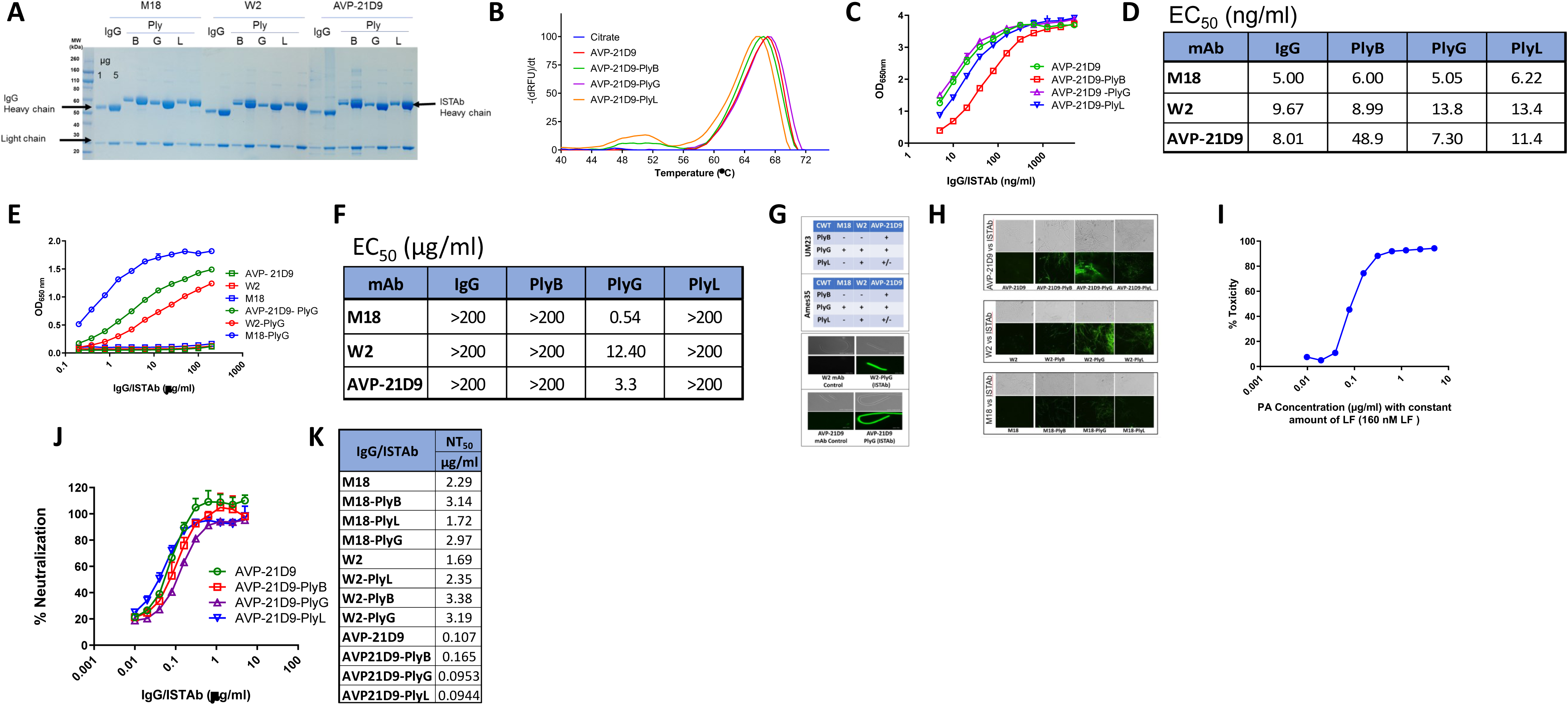
Characterization of ISTAbs. **(A)** SDS-PAGE analysis of ISTAbs and parental IgGs. MW designates molecular weight markers. Positions of IgG and ISTAb are indicated by arrows. Each protein was run in two different concentrations: 1 and 5 µg per lane (as labeled in the first two lanes). **(B)** Thermal unfolding profile of AVP-21D9 IgG and its ISTAbs indicating equivalent thermal stability. **(C)** ELISA measurement of dose-dependent binding of AVP-21D9 and its ISTAb derivatives to PA. **(D)** EC_50_ (ng/ml) table for PA binding of nine ISTAbs and three parental IgGs. **(E)** Whole-cell ELISA for binding to *B. anthracis* strain UM23. Detection was with anti-human IgG conjugate. **(F)** EC_50_ (µg/ml) table summarizing the binding abilities of the nine ISTAbs and three parental IgGs to *B. anthracis* UM23 strain. **(G)** The ability of candidate ISTAbs to bind attenuated B. anthracis strains (BSL2) was analyzed by immunofluorescence microscopy and qualitative data are shown (top). Fluorescent images (bottom) show examples of parental IgG versus ISTAb binding for W2 and AVP-21D9. **(H)** ISTAb binding to *B. anthracis* Ames strain (BSL3) is shown. *B. anthracis* bacteria were incubated with each ISTAb to allow binding and were visualized by fluorescent microscopy after the addition of a secondary antibody against the ISTAb. Phase contrast (top) and fluorescent images (bottom) are shown. **(I)** CellTiter Glo assay of dose-dependent toxicity of 160 nM LF combined with PA toxin. **(J)** Anthrax toxin neutralization by AVP-21D9 ISTAbs and parental IgG. **(K)** NT_50_ values in µg/ml for the three IgGs and their nine corresponding ISTAbs.

### ISTAb and parental IgG binding to PA and B. anthracis

We performed ELISA to measure binding of the nine ISTAbs and the parental IgGs to PA and to whole bacterial cells. Except for AVP-21D9-PlyB, binding of ISTAbs and parental IgGs to PA were indistinguishable, indicating that the CWT did not affect antigen binding (Fig. 3C and D). As expected, parental IgGs did not bind to the surface of *B. anthracis* (Fig. 3E and F). All three ISTAbs containing the PlyG CWT bound the *B. anthracis* surface in whole cell ELISA, whereas ISTAbs containing PlyB or PlyL CWTs showed high EC_50_ (>200 µg/ml) in this assay (Fig. 3F). These data confirm that both Fab and CWTs are functional in the PlyG-based ISTAbs.

### Microscopic visualization of ISTAb binding to the bacterial surface

All nine *B. anthracis* ISTAbs were evaluated for binding to the bacterial surface, utilizing an AlexaFluor 488-labeled mouse anti-human IgG1 Fc secondary antibody for microscopic visualization. The AVP-21D9 based ISTAbs bound attenuated (BSL-2) strains of *B. anthracis* UM23 and Ames35 regardless of the origin of the CWT domain (Fig. 3G). In contrast, the W2-based ISTAb did not bind UM23 or Ames35 strains when the PlyB CWT was utilized, and the only M18-based ISTAb that bound *B. anthracis* contained the PlyG CWT. Taken together, the PlyG CWT is clearly the best candidate for construction of ISTAbs as it bound both *B. anthracis* UM23 and Ames35 strains with all three mAb scaffolds. Similarly, when ISTAbs were tested against the fully virulent Ames strain of *B. anthracis*, the parental IgGs did not show any binding whereas the PlyG CWT-based ISTAbs showed the strongest binding (Fig. 3H). Importantly, the lead ISTAb candidate (AVP-21D9-PlyG) also binds capsulated *B. anthracis* Ames strain (Fig. S1), indicating that the capsule does not interfere with the function of ISTAbs.

### B. anthracis ISTAb toxin neutralization and binding affinities

We next determined whether ISTAbs retain the neutralizing activity of the parental antibody using a cell viability assay in presence of purified anthrax toxin (Fig. 3I). Dose response toxin neutralization plots for AVP-21D9 and its respective ISTAbs demonstrate that the ISTAbs retained neutralization activity similar to parental IgGs (Fig. 3J), with AVP-21D9-derived ISTAbs exhibiting >10 times better neutralization than M18 or W2 ISTAbs (Fig. 3K).

We also measured the affinity of PlyG-containing ISTAbs for PA using Biolayer Interferometry (Fig. S2A-D). The apparent *K_D_* values for W2, W2-PlyG, AVP-21D9, and AVP-21D9-PlyG binding to PA were 0.5, 0.6, 0.001, and 0.06 nM, respectively. The reduction in the affinity of AVP-21D9-PlyG compared to parental IgG was a result of faster off-rate; however, the ISTAb still maintained picomolar affinity for PA (Table 1, Fig. S2A-D), consistent with the neutralizing activity (Fig. 3J and K).

**Table 1.** A summary of *K*_D_ values for binding to PA.

| Sample | $K_D$ (nM) | $K_{on}$ (1/Ms) | $K_{off}$ (1/s) | $R^2$ |
| --- | --- | --- | --- | --- |
| W2 | $0.5 \pm 0.03$ | $1.7 \pm 0.008 \times 10^5$ | $8.9 \pm 0.6 \times 10^{-5}$ | 0.9953 |
| W2-PlyG | $0.6 \pm 0.03$ | $1.8 \pm 0.008 \times 10^5$ | $9.9 \pm 0.5 \times 10^{-5}$ | 0.9961 |
| AVP-21D9 | $0.001 \pm 0.03$ | $2.3 \pm 0.01 \times 10^5$ | $< 0.01 \times 10^{-5}$ | 0.9923 |
| AVP-21D9-PlyG | $0.06 \pm 0.002$ | $2.5 \pm 0.01 \times 10^5$ | $1.6 \pm 0.6 \times 10^{-5}$ | 0.9927 |

### Binding to Fcγ receptors

To ensure that fusion of IgG with the PlyG CWT did not interfere with FcR binding on the surface of innate immune cells, we measured the binding affinity of AVP-21D9 versus the derivative PlyG ISTAb for multiple FcγR proteins. Both AVP-21D9 and the PlyG ISTAb showed strong binding to FcγR1A, FcγR2A, FcγR2B, and FcRN (Fig. 4A; Supplemental Table S2 and S3). There was no notable difference in the binding affinities of different FcR proteins for AVP-21D9 compared to AVP-21D9-PlyG. These data clearly indicate that the CWTs in the ISTAb fusion protein do not interfere with the FcγR binding, similar to our findings for PA binding.

**Fig. 4:**
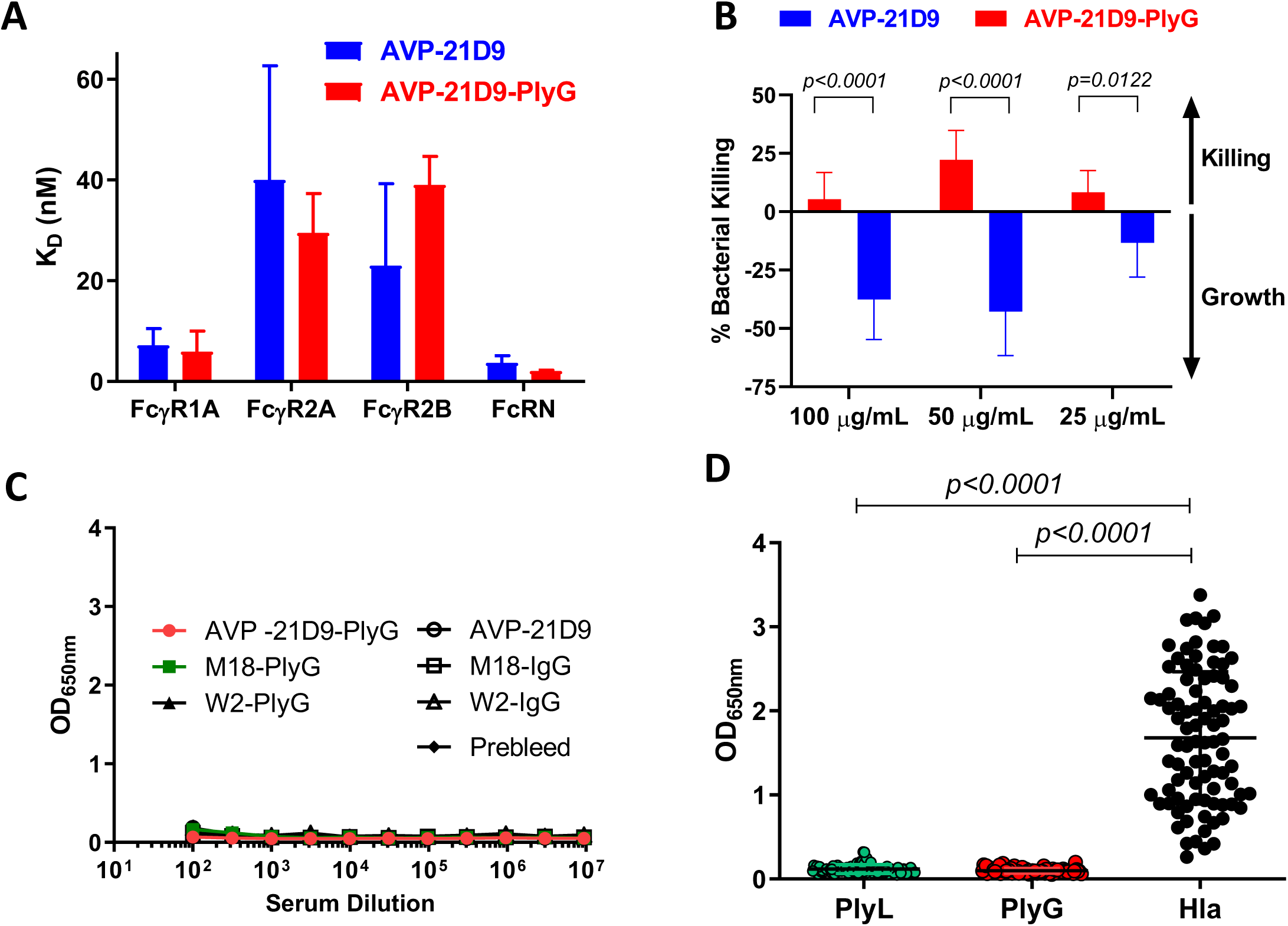
Binding to Fcγ receptors, OPK, Immunogenicity, and seroprevalence studies of ISTAbs. **(A)** Dissociation constants for binding of AVP-21D9 and AVP-21D9-PlyG to various Fcγ receptors. Each bar represents an average of three independent experiments. **(B)** Opsonophagocytic activities of AVP-21D9 versus AVP-21D9-PlyG at 100, 50, and 25 µg/ml using HL-60 cells. Columns represent the average values at a 95% confidence interval (CI) for 33 data points from four independent experiments. **(C)** Immunogenicity measurement of CWT domain of ISTAb derivatives of M18, W2, and AVP-21D9 in comparison with respective IgGs as determined by ELISA, using plates coated with isolated PlyG domain. Samples were run in duplicate. **(D)** Seroprevalence of isolated PlyL and PlyG CWT domains among healthy humans. The reactivity of 88 human sera from healthy donors was tested by ELISA at 1:300 dilution. *S. aureus* Hla was used as a control antigen. Data were analyzed by unpaired Student’s t-test.

### B. anthracis ISTAb opsonophagocytic activities

We analyzed the OPK of the AVP-21D9 versus AVP-21D9-PlyG using HL-60 cells differentiated to neutrophils. HL-60 cells were incubated with three different concentrations of AVP-21D9 or the corresponding ISTAb derivative AVP-21D9-PlyG, serum-containing complement, and *B. anthracis* UM23, for 30 min at room temperature (RT) followed by 45 min at 37°C, and the bacteria were then enumerated. As shown in Fig. 4B, bacterial killing was observed only with the ISTAb while bacteria displayed growth only with the parental antibody, indicating that the ISTAb can mediate opsonophagocytic activity.

### Immunogenicity towards the B. anthracis ISTAb CWT

Antibody responses, if elicited towards the CWT domain of ISTAbs, could potentially reduce the therapeutic efficacy. In order to examine potential immunogenicity of the PlyG CWT domain, we treated mice with PlyG-ISTAb or parental IgG for AVP-21D9, M18, and W2, and measured serum antibody titers against the PlyG CWT. As shown in Fig. 4C, no significant binding was observed, indicating a lack of immunogenicity towards the PlyG CWT.

### Seroprevalence of phage CWT domains in healthy humans

While lack of immunogenicity in mice is reassuring, a potential concern for therapeutic proteins fused with a CWT would be pre-existing human antibodies to CWTs that could inactivate it by blocking CWT binding to the bacterial cell wall. To address this concern, we screened 88 healthy human plasma samples for antibody titers against CWTs for PlyL and PlyG. As a positive control, we used the staphylococcal alpha toxin (Hla), an antigen to which many people have been exposed (*34*). As expected, we observed high titers against Hla, but nearly undetectable titers against PlyL and PlyG (Fig. 4D) indicating low seroprevalence against these two CWTs.

### Stability of ISTAbs

AVP-21D9 and AVP-21D9-PlyG were stored at 1 mg/ml at 4°C, 25°C, or 40°C and tested after 1 month by SDS-PAGE, size exclusion HPLC, and toxin neutralization assay (TNA) to determine the structural and functional integrity of the proteins. As the SDS-PAGE and HPLC data for the 1-month time point demonstrate (Fig. S3A and B) both the parental IgG and its derivative ISTAb remained remarkably stable over the study period. The samples also retained functional activity by TNA assays (Fig. S3C). As isomerization of mAbs is a critical productibility issue, we carried out LC-MS on AVP-21D9 and AVP-21D9-PlyG stored at 40°C for 1 wk. Based on the data, no significant risk was reported for iso-Asp deamidation (Fig. S3D).

### Pre-challenge mouse efficacy studies for subcutaneous (s.c.) route of infection

We first determined the sub-protective doses for the two parental antibodies, AVP-21D9 and W2, in order to determine a range where potential improvements by their derivative ISTAbs could be detected. Groups of five Swiss Webster mice (8 weeks old) were injected s.c. with the antibodies, or buffer control, two hours before s.c. challenge with 10LD_50_ of *B. anthracis* Ames. Health scores were recorded daily for 14 days. The results demonstrated 40% survival in mice that received W2 at 20 μg/mouse or AVP-21D9 at 15 μg/mouse respectively (Fig. 5A and B). These partial rescue doses were selected for pre-challenge evaluation of the ISTAbs (AVP-21D9-PlyG and W2-PlyG). As above, mice were administered with the parental antibodies (AVP-21D9 at 15 μg/mouse, or W2 at 20 μg/mouse; n=10 per group), or an equimolar amount of the derivative ISTAbs, and challenged two hours later s.c. with 10 LD_50_ *B. anthracis* Ames. As shown in Fig. 5C and D, both ISTAbs provided a higher level of survival compared to the parental IgGs. Residual bacterial burden in the surviving animals treated with either AVP-21D9 or AVP-21D9-PlyG was also determined. While not statistically significant, perhaps due to the small number of remaining mice, a clear trend was observed toward lower bacterial burden in the liver, lungs, and spleen of the ISTAb-treated animals compared to the IgG group (Fig. 5E). None of the surviving animals had detectable bacteria in their blood at the study termination.

**Fig. 5:**
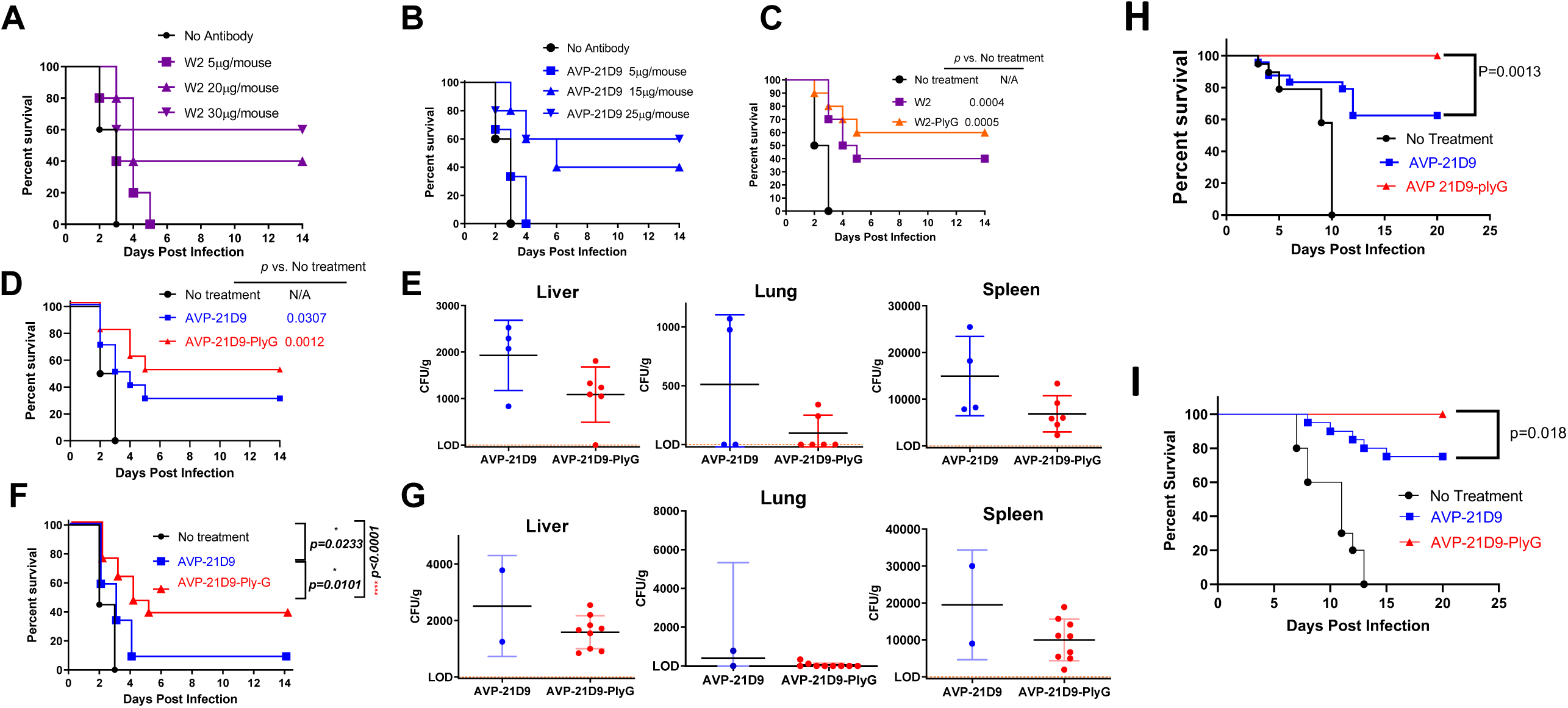
Mouse efficacy studies. **(A,B)** Sub-protective dose determination for AVP-21D9 and W2 antibodies. Mice were injected with varying doses of either AVP-21D9 or W2 IgGs, or with vehicle control, prior to s.c. challenge with the Ames strain of *B. anthracis*. Mice were monitored daily for morbidity and mortality for a period of 14 days post-challenge. **(C, D)** Efficacy of the parental IgGs or their derivative ISTAbs delivered two hours before s.c. infection with Ames spores. Statistical analysis was performed using the Log-rank (Mantel-Cox) method. **(E)** Quantitation of remaining bacterial load (CFU/g) in liver, lung, and spleen of surviving mice from the pre-exposure studies. **(F)** Efficacy of 330 µg/mouse of AVP-21D9, or a molar equivalent amount of its derivative ISTAb (AVP-21D9-PlyG), delivered 18-24h post-infection with Ames spores using the s.c. route of challenge. Statistical analysis was performed using the Log-rank (Mantel-Cox) method. **(G)** Quantitation of remaining bacterial load (CFU/g) in liver, lung, and spleen of surviving mice from the post-exposure studies on day 14 post-challenge. **(H)** Pre-challenge mouse efficacy of AVP-21D9 and AVP-21D9-PlyG in respiratory infection model of anthrax. **(I)** Post-challenge mouse efficacy of AVP-21D9 and AVP-21D9-PlyG in respiratory infection model of anthrax. Mice were challenged intranasally with the Ames strain and 48 hours post-challenge were treated via the IP route with either AVP-21D9 (195 µg/mouse) or the molar equivalent of AVP-21D9-PlyG (216 ug/mouse), or with vehicle control. Mice were monitored daily for morbidity and mortality for a period of 20 days post-challenge.

### Post-challenge mouse efficacy studies for s.c. route of infection

The post-challenge therapeutic efficacy of AVP-21D9-PlyG compared with parental AVP-21D9 antibody was determined in two independent experiments, with delay of treatment to 18-24 hr post-challenge. A higher IgG dose of 330 µg/mouse and an equimolar ISTAb dose was used in these studies. The combined data of these two studies (n=24 for treatment group, and n=20 for control) are shown in Fig. 5F. Statistical analysis (Log-rank) showed highly significant protection provided by AVP-21D9-PlyG compared to the untreated group (p<0.0001). Furthermore, the difference in protection provided by the ISTAb *vs*. parental AVP-21D9 was also statistically significant (p=0.0101). Bacterial burden in the liver, lung, and spleen of surviving animals was consistently lower in ISTAb-treated animals compared to parental IgG (Fig. 5G).

### Pre-challenge mouse efficacy studies for respiratory route of infection

Pre-challenge efficacy of the AVP-21D9-PlyG against respiratory *B. anthracis* challenge was compared with its parental IgG, AVP-21D9. Mice received intraperitoneal (i.p.) treatment with either buffer vehicle as control (n=19), or molar-equivalent IgG or ISTAb doses of 65 µg/mouse and 72 µg/mouse respectively (n=24 per group). At two hours post-treatment, the mice were challenged intranasally (i.n.) with 5LD_50_ *B. anthracis* Ames spores and monitored daily for morbidity and mortality for 20 days (Fig. 5H). All surviving mice appeared completely healthy based on the scoring criteria, and statistical analysis (Log-rank) showed highly significant protection provided by AVP-21D9-PlyG (100% survival) compared to the untreated group (p<0.0001). Furthermore, the statistical difference in protection provided by the ISTAb vs. parental AVP-21D9 was also highly significant (p=0.0013).

### Post-challenge mouse efficacy studies for respiratory route of infection

In two fully independent experiments, AVP-21D9-PlyG was compared with parental IgG for therapeutic efficacy against the respiratory route of anthrax infection. Mice were challenged i.n. with 5LD_50_ *B. anthracis* Ames. Then, at 48 hr post-challenge, the mice received i.p. administration of either vehicle as control (n=10), or molar-equivalent IgG or ISTAb doses of 195 µg/mouse and 216 µg/mouse respectively (n=20 per group). The mice were monitored daily for morbidity and mortality for a period of 20 days post-challenge. Combined data for these two studies are shown in Fig. 5I. All surviving mice at the end of both studies appeared completely healthy based on scoring criteria, and statistical analysis (Log-rank) showed highly significant protection provided by AVP-21D9-PlyG (100% survival) compared to the untreated group (p<0.0001). Furthermore, the difference in protection provided by the ISTAb vs. parental AVP-21D9 was also statistically significant (p=0.0182). These data, together with the post-challenge results for the s.c. route of infection, establish the proof-of-concept that conferring surface binding property to a neutralizing PA antibody significantly enhances its *in vivo* therapeutic efficacy.

### Pharmacokinetic analysis of B. anthracis ISTAbs in mice

As a preliminary evaluation of the relative bioavailability of ISTAbs versus IgG, we treated a group of three mice with AVP-21D9, M18, W2, or molar equivalents of their respective PlyG ISTAbs. Samples were collected on days 3, 7, 14, 21, and 28. IgG levels (µg/ml) were measured using a quantitative sandwich ELISA. As shown in Fig. 6A-C, serum antibody concentrations were higher in the IgG groups on day 3 compared to ISTAbs. This is remarkable in that the ISTAbs showed superior efficacy despite apparent lower bioavailability compared to parental antibodies.

**Fig. 6:**
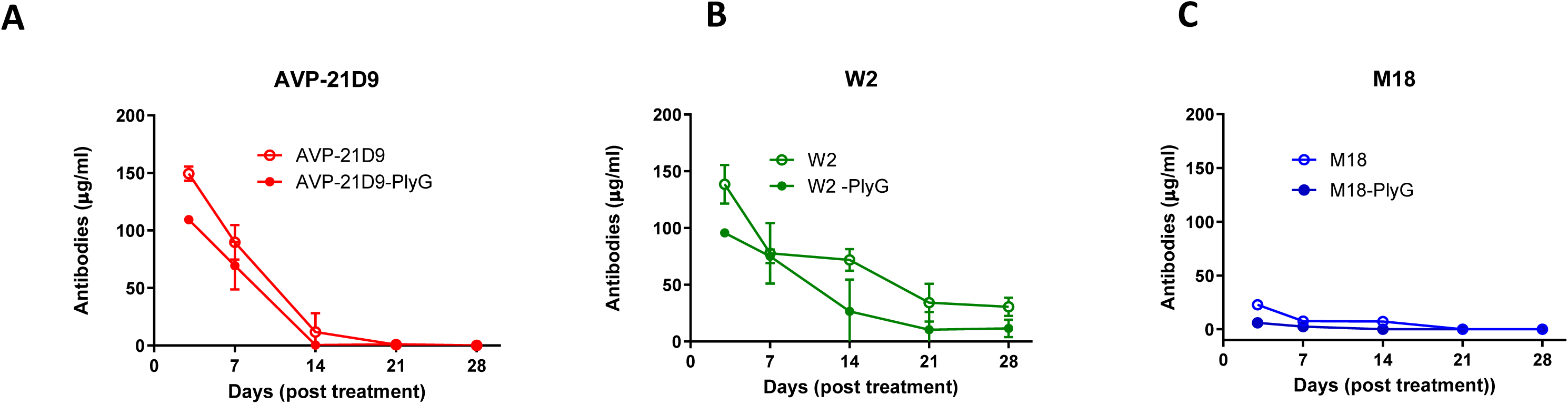
Pharmacokinetic studies in Mice. The pK measurement of CWT domain of ISTAb derivatives of AVP-21D9, W2, and M18 in comparison with respective IgGs as determined by ELISA, using plates coated with isolated PlyG domain. Samples were run in duplicates. **(A-C)** Preliminary pK analysis of ISTAb vs. IgG. Mice (n=3) were treated with 330 µg/animal of either IgG or ISTAb on day 0. Serum samples were collected on days 3, 7, 14, 21, and 28 to compare the anti-PA binding titers. Mean values from three mice and associated errors are plotted

### ISTAb evaluation in a rhesus macaques inhalational anthrax model

We performed a therapeutic evaluation of ISTAb with separate cohorts (n=5, AVP-21D9; n=5, AVP-21D9-PlyG; n=4, sham-treated controls), with all groups receiving equivalent comparable therapeutic or sham doses (20 mg/kg) by intravenous administration 24h post aerosol challenge. Initially, all animals were aerosol challenged with a target inhalation dose of 1.0E+08 of virulent *B. anthracis* spores (Ames). Results of the aerosol challenge (Fig. 7A) shows individual dosing was not significantly different between experimental groups, and the overall mean delivered dose (3.32E+08 ± 1.73E+08) approximated the target challenge dose. All animals in AVP-21D9 and AVP-21D9-PlyG treatment groups survived lethal aerosol challenge, whereas sham-treated animals succumbed to disease 112-128h post challenge (Fig. 7B). All four animals in sham-treatment control group demonstrated blood bacteremia (Fig. 7C) indicative of hematogenous spread and fulminant anthrax disease, including prominent bacterial loading in bronchoalveolar lavage (BAL) samples taken at necropsy (Fig. 7D). No animals in either AVP-21D9 or AVP-21D9-PlyG treatment groups experienced blood bacteremia and survived until termination of the experiment (day-21 PI). Residual bacterial loading in necropsy BAL samples in the AVP-21D9 and AVP-21D9-PlyG groups were observed (Fig. 7D). Colony counts in the lung lavage of treated animals may have been the result of ungerminated spores sequestered in the lung at day 21 post challenge.

**Fig. 7:**
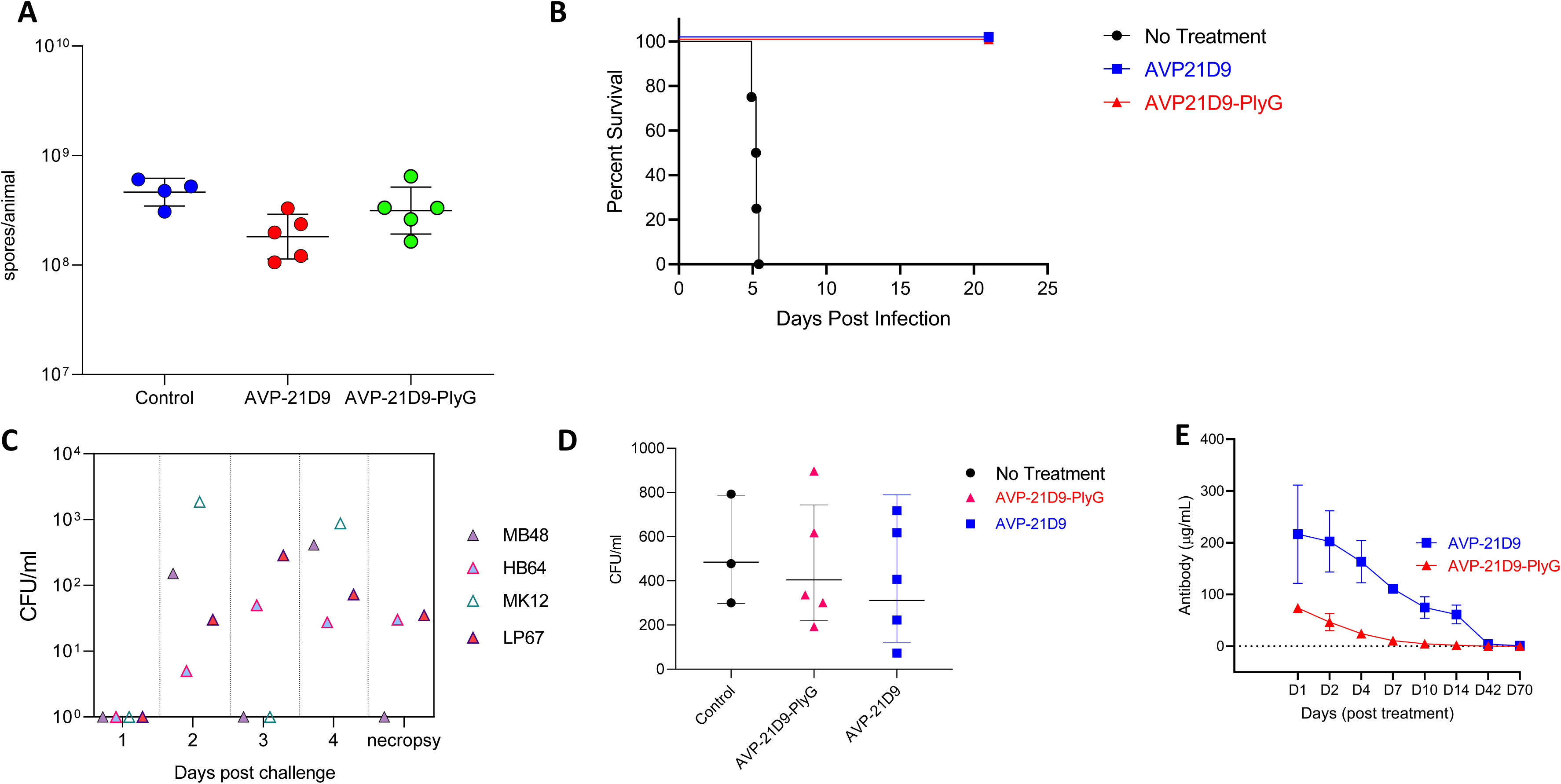
Nonhuman primate efficacy studies. Rhesus macaques (n=4) were exposed by aerosol to *B. anthracis* sp. Ames. **(A)** Determination of individual animal aerosol inhaled challenge dose (CFU/animal), **(B)** Survival curve, **(C)** Blood bacteremia in control animals postexposure excluding one animal (MK12) where samples were impossible to obtain; there was no detectable bacteremia in any of the treated animals during any times postexposure, including necropsy, **(D)** Determination of bacterial concentrations from bronchoalveolar lavage samples taken at necropsy. **(E)** NHP pK study. NHPs (n=4) were infused with 20 mg/kg of either AVP-21D9-PlyG or AVP-21D9-IgG and the concentration of antibodies in serum was measured at days D1, D2, D7, D10, D14, D42, and D70. Mean values of concentration of antibody from four NHPs and error bars for standard deviations were plotted.

### Pharmacokinetic studies in NHPs

Serum samples from two male and two female rhesus macaques treated by IV infusion with a single dose (20 mg/kg) of AVP-21D9 or AVP-21D9-PlyG were collected after days 0, 1, 2, 4, 7, 10, 14, 42, 70, and 99. As shown in Supplemental Table S4, t1/2(d) and C_max_ (μg/ml) values of AVP-21D9-PlyG were 4.21 (days) and 73.88 (μg/mL) respectively, which were two folds and three folds lower than the t1/2(d) and C_max_ (μg/ml) of parental IgG, respectively. The serum concentration plot (Fig. 7E) exhibited a significantly lower bioavailability of the PlyG ISTAb when compared with parental IgG (multiple t-test, p=<0.001). These data further demonstrate the efficacy of ISTAbs despite suboptimal PK properties.

### Immunogenicity studies

NHP PK serum samples from days 42, 70, and 99 were analyzed for anti-PlyG antibodies, especially in group 1 NHP, which were treated with 20 mg/kg of AVP-21D9-PlyG. As shown in Fig. S4, there were no significant anti-PlyG titers detected in group 1 NHP treated with AVP-21D9-PlyG (ED_50_ <1.0).

### Application of ISTAb technology to C. difficile

As a proof of concept for broad applicability of the technology, we also generated and characterized several ISTAb for *C. difficile* as another Gram-positive pathogen. We bioinformatically screened all sequenced *C. difficile* bacteriophages as well as all sequenced *C. difficile* bacterial genomes, which contain prophage sequences, for potential phage endolysins. Twenty-five endolysin or endolysin-like sequences were analyzed using the PFAM database to identify catalytic domains and define domain boundaries. Endolysins that lacked known CWT domains or had less than 50 non-catalytic domain amino acids, were excluded from further analysis. The remaining CWTs were aligned and compared for percent sequence identity. Duplicate sequences were eliminated and those with >95% identity was condensed into one archetype sequence. Eventually, six *C. difficile* endolysin CWT families (630A, QCD, CD27L, C2, CD119, and CCL2), which ranged from 30% to 90% identity, were selected for further study (Fig. S5A). All six *C. difficile* CWTs were chemically synthesized, expressed, and qualitatively screened for binding by fluorescent microscopy against 11 *C. difficile* strains and 12 additional anaerobic strains that included those known to colonize the gastrointestinal tract (i.e., other members of the Clostridiaceae, Lactobacillaceae, and Bacteroidaceae families). Ultimately, the 630A and CCL2 CWTs were selected for generation of *C. difficile* ISTAbs because they displayed: 1) high yield and ease of purification (Fig. S5B); 2) the ability to bind all *C. difficile* strains with negligible or no binding to non-*C. difficile* strains (Fig. S5C); and 3) the lowest homology between all six CWTs (30% identity, Fig. S5A). A representative image showing binding of AlexaFlour-labeled 630A to *C. difficile* strain BAA1812 is shown in Fig. S5D).

For ISTAbs generation for *C. difficile*, we selected the well characterized humanized monoclonal anti-TcdA (6G5) and anti-TcdB (9H10), and the 630A (accession no. YP_001088405.1) and CCL2 (accession no. CCL24417.1) as CWTs. We characterized both IgGs and ISTAbs using SDS-PAGE (Fig. S5E), size exclusion HPLC (Fig. S5F), and binding and neutralizing activity assays. The binding curves for parental antibodies and ISTAbs were essentially indistinguishable (Fig. S5G). Using a Caspase-based cytotoxicity assay (*35*) for TcdA and TcdB (Fig. S5H) we showed that both IgGs and ISTAb exhibit comparable neutralizing potency (Fig. S5I-K). All four ISTAb constructs possessed binding specificity for *C. difficile* with little to no binding observed for non-*C. difficile* strains or species (Fig. S5C). Furthermore, all 630A ISTAbs consistently showed strong binding to FcγR1A, R2A, R3A and low binding to the inhibitory FcγR2B for both TcdA (Fig. S6A) and TcdB (Fig. S6B). In no instance was the binding to the respective receptor lower for ISTAbs compared to parental antibodies.

We also analyzed sero-reactivity of 47 human serum samples to the *C. difficile* CCL2 and 630A CWTs. As controls, we used the *S. aureus* toxic shock syndrome toxin 1 (TSST-1), for which we expect seroprevalence, and purified glycoprotein (GP) of Ebola virus as negative control. As shown in Fig. S6C, while we observed strong reactivity of the sera to TSST-1 and a few serum samples unexpectedly showed some reactivity to Ebola GP, the lowest reactivity was observed for 630A and CCL2 (significantly lower than TSST-1, p<0.0001). The reactivity of 630A, but not CCL2, was significantly lower than Ebola GP (P=0.0202). Stability studies also showed that the IgG and ISTAbs remained remarkably stable over the study period (Fig. S7A-C).

### C. difficile ISTAb in vivo efficacy study in Hamsters

Prior to spore challenge, Golden Syrian hamsters were treated on days -1, 0, +1, and +2 relative to infection (similar to regimen reported previously for bezlotoxumab) with either parental IgG or molar equivalent of a cocktail of TcdA and TcdB ISTAbs (1 mg/hamster per day). On Day -1, animals received a single dose of 10 mg/kg clindamycin by intraperitoneal injection (IP) to wipe out the intestinal flora. On day 0, animals received an intragastric dose of 100 spores of *C. difficile* clinical isolate 630 (ATCC^®^ BAA-1382™). While there were no survivors in the group treated with control IgG, the cocktail treatment group showed 60% survival (Fig. S6D) and the weight loss started to level off about 4-5 days post infection (Fig. S6E).

## Discussion

Engineered antibodies that embody multiple functions into a single molecule are highly desirable therapeutics. Recent advances in antibody engineering allow for the generation of antibodies consisting of two different heavy/light chain combinations, or multiple antigen binding domains, each possessing distinct binding specificities. These “bispecific” mAbs can simultaneously bind two different antigens or epitopes of the same antigen to increase affinity or enable bifunctionality. As an example, a bispecific mAb (BiS4Pa) was created that targets Psl, the *Pseudomonas aeruginosa* exopolysaccharide, and PcrV, a type III secretion system protein, and demonstrated superior protection in a *P. aeruginosa*–induced murine pneumonia model compared to parental mAb combinations (*36*). In another study, an antibody-antibiotic conjugate was used to kill intracellular *S. aureus* (*37*). In this approach, an IgG bound the conserved *S. aureus* wall teichoic acid and upon Fc-mediated OPK phagolysosomal proteases cleaved a labile bond releasing rifampicin conjugated to the mAb. This antibody-antibiotic conjugate was shown to be superior to vancomycin *in vitro* and *in vivo.* Our current study expands the concept of bifunctional antibodies to integration of toxin neutralization and Fc-mediated effector functions into a single molecule in form of ISTAbs.

ISTAbs have several favorable features as shown in Fig.1B and C: *i)* They show specific and high-affinity binding to the infectious agent, mediated by the CWT (*38*); *ii)* They are expected to enhance binding by the avidity effect since each ISTAb carries two CWT domains; *iii)* They are expected to accumulate on the bacterial surface at the site of infection where the anti-toxin is needed; *iv)* They are expected to sequester toxin to prevent release in the tissues and circulation; and, *v)* ISTAbs provide opsonic activity to enhance clearance of the bacteria and targeted toxin by the phagocytes that engulf the bacteria.

As proof of concept, we have tested ISTAb efficacy against both *B. anthracis* and *C. difficile* infections. Currently, there is no approved vaccine for C. difficile, and the only approved *B. anthracis* vaccine is plagued with reactogenicity and other side effects. Notably, challenges to vaccination strategies that stem from the inability to generate a long-lasting and protective response continue to exist, as manifested by very recent clinical trial failures of both Sanofi and Pfizer *C. difficile* vaccines (*39*). Immunotherapeutics as agents that can be used post-expodure and provide immediate protection both in general population and immunosuppressed patients remain an important tool for management of bacterial infections.

PA has been a key target for anthrax immunotherapies (*4-6*). A fully human mAb, called raxibacumab, is the first biological product to be approved through the FDA Animal Rule (*40*), and shown to be effective in New Zealand white rabbits and cynomolgus macaques. Raxibacumab was found to be safe at 40 mg/kg in clinical studies with more than 300 healthy adults, including 43 individuals receiving a double dose (*41*). Another human mAb, called MDX-1303 (Valortim), was shown to be effective in prophylactic and post-symptomatic treatment of rabbits exposed to aerosolized *B. anthracis* spores, and a single intramuscular injection of 1 mg/kg of body weight fully protected monkeys challenged with aerosolized *B. anthracis* spores (*42*). Based on the published data, the IC_50_ of raxibacumab is 0.21 nM (*43*), which is very close to the lead IgG molecule AVP-21D9 (IC_50_ = 0.3 nM) used in our study, and we have shown that the AVP-21D9-PlyG ISTAb has significantly better protection than AVP-21D9 in post-exposure studies of anthrax challenge using a mouse model. Therefore, we expect that AVP-21D9-PlyG will have superior protection compared to raxibacumab. The key advantage of our approach over raxibacumab is that ISTAbs display the same anti-toxin properties while at the same time they bind to the surface of the bacteria to trigger an innate immune response to clear the infection. This rapid clearance of bacteria is expected to significantly enhance the efficacy of the ISTAbs beyond what has been achieved with raxibacumab. In support of superior ISTAb effects, results similar to *B. anthracis* studies were obtained with our *in vitro* and *in vivo* analysis of ISTAb efficacy against *C. difficile*. In addition to inducing bacterial clearance by triggering OPK and toxin neutralization, ISTAbs create a concentration gradient towards the site of infection, where these therapeutic molecules are needed the most. A similar class of therapeutic molecules, called lysibodies, contain an IgG Fc fused with an endolysin CWT domain targeting *S. aureus* (*44, 45*). Lysibodies were shown to promote phagocytosis by macrophages and neutrophils, and to protect mice from methicillin-resistant *S. aureus* (MRSA) infection in two model systems (*44, 45*). Unlike ISTAbs, lysibodies do not possess the Fab fragment of IgG (*44*), so they are devoid of antigen specific binding and/or neutralization. In this respect, ISTAbs represent a superior therapeutic by combining surface binding with toxin neutralization, particularly for pathogens that utilize secreted virulence factors such as toxins.

Although the core IgG of the AVP-21D9-PlyG ISTAb is of human origin, it could be a potential drawback if the PlyG CWT domain leads to immunogenicity, as anti-CWT antibodies could neutralize these therapeutic molecules. However, treatment with an ISTAb would likely be used only a few times in a patient’s lifetime when a life-threating infection occurs, so the immunogenic potential of CWT would be a minor concern. Moreover, our studies clearly addressed this issue showing that the CWT domains, specifically PlyG, is not immunogenic in a mouse model when ISTAb administration mimicked the expected clinical treatment dose and route (Fig.4C). Additionally, naturally occurring antibodies against the PlyG and PlyL CWTs were not detected when plasma from 88 individuals were analyzed (Fig. 4D) for preexisting antibodies titers. Consistent with these findings, we obtained the same results for the CWT domains of the ISTAbs used in the *C. difficile* studies. Moreover, there is evidence from multiple endolysins suggesting that even if antibodies are produced against these proteins, they are non-neutralizing, presumably due to the high-affinity of the CWTs for the bacterial surface (*46, 47*).

The observed efficacy of ISTAbs is remarkable in light of our data showing that they have lower bioavailability. This suggests that further engineering of these molecules to improve PK properties can further increase efficacy of ISTAbs. Thus, future direction involves introduction of mutations in the Fc domain of ISTAbs such as YTE and LS mutations that increase IgG’s affinity for neonatal Fc receptor at acidic pH and promote recycling of IgG back into circulation.

In summary, we tested and evaluated the ISTAbs as a new therapeutic platform technology, which could target any secreted virulence factor (toxins, enzymes, and other immunomodulating proteins) on the bacterial surface at the site of infection. Using anti-toxin ISTAbs, we have shown that ISTAbs maintain whole cell binding as well as toxin neutralizing functions. ISTAbs also trigger OPK in contrast to the parental IgGs. Most importantly, the anti-PA AVP-21D9-PlyG ISTAb confers significantly better protection when efficacy is compared with equimolar IgG alone in a post-challenge anthrax mouse model. These observations provide proof of principle for wide ranging applications of this technology against toxigenic bacteria.

## Methods

### Identification of CWTs for ISTAb construction

Phylogenetic trees based on seed alignments of all accumulated *B. anthracis* endolysins were assembled by FastTree on 100 resamples. Five unique *B. anthracis* endolysins were ultimately selected for gene synthesis. These include the CWT domains from PlyG (ABB55421.1), the endolysin of the γ phage, PlyB (YP_009031336), the endolysin from the Bcp1 phage, PlyL (NP_846312.1), the endolysin from the lambda B02 prophage lysogenized in the *B. anthracis* Ames strain, PlyAP50 (YP_002302543.1), the putative endolysin from the AP50 phage, and PlyTsamsa (YP_008873459.1), the endolysin from phage Tsamsa. *C. difficile* specific CWT identification method has been described in Supplemental Method 1.

### Expression and purification of CWTs

Five endolysin CWT domains from *B. anthracis*-specific phages and six from *C. difficile* were codon-optimized for expression in *E. coli*, synthesized (GeneArt), and subcloned into an arabinose-inducible pBAD24 expression vector, sequenced (Macrogen) to confirm identity, and eventually transformed into BL21 (DE3) competent *E. coli.* Proteins were expressed in *E. coli* and cell lysates were used to extract and purify CWTs. An example figure for the PlyL and PlyG CWTs based on pBAD24 expression is shown in Fig. S8.

### Bacterial binding specificity of CWTs

The PlyL and PlyG CWTs (*B. anthracis*), and CCL2 and 630A (*C. difficile*), were chemically crosslinked to AlexaFluor^®^-555 according to manufacturer’s instructions (ThermoFisher Scientific). Briefly, 0.5 ml of CWTs (2.0 mg/ml) was mixed with 50 µl of 1 M sodium bicarbonate and reacted with 20 µl of AlexaFluor^®^-555 dye (2 mg/ml in DMSO). The reaction mixture was incubated at RT for 1 hr and unreacted dye was removed by application to a PD-10 desalting column (GE Healthcare).

*Bacillus* strains (Fig. 2B and 2C) and *C. difficile* strains (Fig. S5C) were obtained from the American Type Culture Collection (ATCC), or BEI Resources, and grown in Brain-Heart Infusion (BHI) medium overnight (o/n). The next morning, cells were washed 2X in PBS and 20 µg of labelled CWT was added to 100 µl of bacterial cells and allowed to incubate at RT for 10 min. The reaction in the absence of fluorescent dye served as a control for autofluorescence. After incubation, labeled bacterial cells were pelleted and washed with PBS, diluted to 100 µl again, and 1 µl of this mixture was applied to a microscope glass slide and sealed with glass coverslip. Bacteria were viewed via both bright field microscopy and fluorescent microscopy by a Nikon Eclipse 80i microscope for all BSL-2 strains or a Nikon Eclipse Ts2 microscope for the BSL-3 strains (*B. anthracis* Ames and ASC506). The NIS-Elements software was utilized for image analysis.

### Seroprevalence of CWT domains in healthy humans

Human serum samples from healthy donors (Emergent BioSolutions plasmapheresis program) (*34*) were assessed for pre-existing antibody titers against the CWTs from PlyL or PlyG, or Hla as a positive control (for *B. anthracis)* and CCL2 and 630A (for *C. difficile*). Proteins (100 ng/well) were coated in NUNC ELISA plates o/n at 4°C. Plates were blocked for 1 h at RT. Serum samples were diluted at 1:200 in PBS and detected using anti-human IgG-HRP. The OD_650nm_ values were recorded and EC_50_ values were analyzed as described previously.

*C. difficile* specific method has been described in Supplemental Method 2.

### Selection and construction of IgGs and ISTAbs

Sequences of three mAbs M18 (accession number 3ETB_F), W2 (ABB02452), and AVP-21D9 (ACK09999), were used to generate variable regions of heavy and light chains in human IgG scaffold. These sequences were cloned into pSF antibody expression vector. CWT domains were cloned with the native linker of the phage endolysin proteins (i.e., sequence linking the catalytic domain with the CWT domain); hence, no external linkers were utilized. All CWT sequences were codon optimized for human cell expression and included amino acids 164-233 of the PlyG endolysin (ABB55421.1), amino acids 191-284 of the PlyB endolysin (YP_009031336), and amino acids 165-234 of the PlyL endolysin (NP_846312.1).

*C. difficile* specific method has been described in Supplemental Method 3.

### Purification and characterization of IgGs and ISTAbs

The ExpiCHO system was used for the production of IgGs and ISTAbs as described previously(*48*). Briefly, one mg of plasmid was transfected into ExpiCHO cells seeded at 6 x10^6^ cells/ml in a 1 L culture medium. Cells were harvested on day 14 and dialyzed against citrate buffer (20 mM citrate + 10 mM glycine + 8% sucrose pH 5.5) after purification.

### Differential Scanning Fluorimetry

Thermal stability of the parental IgGs and ISTAbs in 1% citrate buffer was assessed using differential scanning fluorimetry (DSF), as described previously (*49*). In summary, SYPRO® Orange dye (Invitrogen, Carlsbad, USA) diluted in sterile dH_2_0 was added to the proteins to reach a final concentration of 6X within a reaction volume of 25 μl in a 96-well hard-shell plate with clear bottom (Bio-Rad). The thermal assay was performed in a Bio-Rad CFX Connect thermocycler wherein fluorescence measurements were made by increasing the temperature from 30 to 99°C at intervals of 0.1°C/6s. The average melting temperature (T_m_) calculated from triplicates was defined as the vertex of the first derivative (dF/dT) of relative fluorescence unit (RFU) values. Bovine serum albumin (Pierce) in 1X Dulbecco’s phosphate-buffered saline (DPBS) pH 7.4 with a T_m_ of 66 ±0.0°C was used as a control.

### ELISA for IgG/ISTAb binding to toxin/bacteria

#### PA/B. anthracis

Standard ELISAs were performed using previously published methods (*50*). Protective antigen (BEI, Manassas, VA) was coated o/n at 4°C in ELISA plates at 100 ng/well. Next day, starting at 5 μg/mL, two folds diluted IgGs and ISTAbs were added after blocking and detected by anti-human IgG-HRP. For whole cell binding assays, NUNC ELISA plates were coated with 100 μL of 0.4% formalin (vol/vol) treated *B. anthracis* UM23 o/n at 4°C. The following day, two-folds diluted ISTAbs and IgGs were used as primary antibodies and detected with goat anti-human IgG-HRP antibody, and OD_650nm_ values were recorded and EC_50_ values determined. *C. difficile* specific method has been described in Supplemental Method 4.

### Octet binding studies

Biolayer interferometry (BLI) kinetic assays were performed on an Octet Red96 System (Fortebio). For kinetic analysis of PA, ligands W2, W2-PlyG, AVP-21D9, or AVP-21D9-PlyG were diluted to 2 µg/ml in 1X kinetics buffer and loaded onto pre-hydrated Protein G sensors (Fortebio) for 120 sec. After loading, a 60 sec baseline was used to establish stable ligand binding before dipping the coated sensors into a serial dilution of PA analyte. Analyte association was maintained for 300 seconds, followed by a 300 second dissociation step. To control for non-specific binding of PA to sensors in the absence of ligand, reference sensors were used in parallel for each concentration of PA and subtracted from total response. For AVP-21D9 and AVP-21D9-PlyG binding kinetics to Fc receptors, recombinant His-tagged FcγR1A, FcγR2A, FcγR2B, or FcRn were loaded onto pre-hydrated Ni-NTA sensors for 120 sec before establishing a steady baseline in 1X kinetics buffer. Coated sensors were then transferred into a concentration range of analyte (AVP-21D9 or AVP-21D9-PlyG) for 300 sec, followed by a 300 sec dissociation step. Experiments were repeated in the absence of Fc receptors to control for non-specific binding. All data analyses were performed with ForteBio Data Analysis 9 software. To obtain *K*_on_, *K*_off_, and *K*_D_ values, specific binding curves were fit globally to a 1:1 Langmuir binding model.

### Toxicity and toxin neutralization assays

Our method was adopted and modified from Nungdi et al. (*51*); *Toxicity assay*: RAW 264.7 cells were allowed to adhere to the plate surface at 37°C, 5% CO_2,_ 95% humidity for 16-18 hr. Two-fold dilutions of PA and LF were prepared starting at 5 µg/ml and added to cells at a 1:1 ratio and incubated at 37°C, 5% CO_2,_ 95% humidity for 3 hours. After incubation, 50 μl of the reconstituted CellTiter-Glo (CTG) reagent was added and luminescence (emission at 560 nm) was measured. Toxicity was calculated as % toxicity = [1 – (% viable cells treated with PA and LF)/ (% viable cells without treatment)] x 100. *Toxin neutralization assay (TNA)*: RAW 264.7 cells were prepared and seeded as described above. Two-fold or semi-log PA and LF dilutions were prepared in 96-well format in the same media, with concentrations starting at 1.25 µg/ml and 0.16 µg/ml, respectively. ISTAbs were diluted to 20 µg/ml. PA and LF were mixed at a 1:1 ratio, and ISTAbs were added at a 1:1 ratio to the PA/LF mix. PA/LF/ISTAb mix was incubated at RT for 30 min. As above, luminescence (emission at 560 nm) was measured after incubation. Toxin neutralization was calculated as % neutralization = (% viable cells treated with IgG or ISTAb and toxin)/ (% viable cells without treatment) x 100.

*C. difficile* specific method has been described in Supplemental Method 5.

### ISTAb/IgG OPK activities

OPK assays were adopted and optimized based on previously described methods (*52-54*). Briefly, HL-60 cells were differentiated into neutrophil-like PMNs by growing in RPMI media containing 1.5% dimethyl sulfoxide (DMSO) and 10% FBS for 7-8 days (differentiation confirmed by expression of CD11b). For the OPK assays, three different final concentrations of 100, 50, and 25 µg/ml of ISTAb or IgGs were made in Opsonization Buffer B (OBB; Hanks’ balanced salt solution (HBSS) supplemented with CaCl_2_, MgCl_2_, 0.5% FBS, and 0.1% gelatin) and added to a sterile 96-well NUNC assay plate (20 μl/well). Stock was diluted to ∼10^5^ CFU/ml and 10 μl/well of this diluted stock was added to the reaction mix. The plate was incubated for 30 minutes in a mini orbital shaker (700 rpm) at RT. Twenty-five microliter of PMN adjusted to 1×10^7^ cells/ml in OBB and 25 µl of 1:8 diluted complement (Baby Rabbit -Pel-Freez Biologicals, LLC) were added to each well (total reaction mix: 80 µl/well). OPK reactions were carried out by incubating for 45 minutes at 37°C, 1% CO_2_. A 10 µl volume of sample was plated on TSB agar plate and bacterial colonies on the agar plates were enumerated. The percent OPK was calculated using the following formula: % OPK = Average CFU count for bacteria without antibody (control) - Average CFU in samples with antibodies (test)/Average CFU count for bacteria without antibody (control) X 100%.

### Mouse ISTAb efficacy studies

#### I. Sub-protective dose optimization

Swiss-Webster mice at 8 weeks of age were injected via s.c route with the antibodies 2 hr before challenge with a lethal dose of the Ames strain of *B. anthracis* spores (10LD_50_). The mice were monitored daily for morbidity and mortality for a period of 14 days post treatment, with daily health score assessments recorded according to our IACUC-approved Animal Health Score Chart.

#### II. Pre-challenge efficacy studies for the s.c. route of infection

Efficacies of selected ISTAbs (AVP-21D9-PlyG and W2-PlyG) against anthrax challenge were compared with their parental antibodies (AVP-21D9 and W2) at the sub-protective doses for AVP-21D9 and W2 that were determined as described above. At 2 hours prior to anthrax challenge, Swiss Webster mice at 8 weeks of age were administered by the s.c. route either the parental antibodies (AVP-21D9 at 15 μg/mouse or W2 at 20 μg/mouse; n=10 per group), or the derivative ISTAbs at equivalent number of moles with respect to their corresponding parental antibodies (n=10 per group). For the subsequent challenge, a lethal dose of *B. anthracis* spores (Ames strain) was administered behind the right foreleg (s.c.,10LD_50_). The following control groups were also included in the study: a) control mice that were infected but received buffer vehicle instead of antibody treatment (n=10); b) control mice that were left uninfected and received buffer vehicle only (n=10). Mice were monitored daily for morbidity and mortality for 14 days post treatment. Daily health score assessments were made according to our IACUC-approved Animal Health Score Chart.

#### III. Post-challenge efficacy studies for the s.c. route of infection

We also tested post-challenge efficacy of AVP-21D9-PlyG in comparison to the parental AVP-21D9 antibody. Swiss Webster mice at 8 weeks of age were first infected with a lethal dose of *B. anthracis* spores (Ames strain) using the s.c. route (5LD_50_). Subsequently, at 18-24 hr post infection, the mice were treated s.c. with either the AVP-21D9 at a dose of 330 μg/mouse or the derivative ISTAb at equivalent number of moles with respect to IgG (n=24 per group). Control mice were treated with buffer vehicle post challenge (n=20).

#### IV. Pre-challenge efficacy studies for the respiratory route of infection

The procedure was the same as described above for the s.c. route of challenge except that a challenge dose of 5LD_50_ was used and the AVP-21D9 at 65 μg/mouse (n=24), or AVP-21D9-PlyG at equivalent number of moles corresponding to 72 μg/mouse (n=24), were administered by i.p. route. Control mice received buffer vehicle prior to infection (n=19).

#### V. Post-challenge efficacy studies for the respiratory route of infection

Swiss Webster mice at 8 weeks of age were first infected with a lethal dose (5LD_50_) of *B. anthracis* spores (Ames strain) via the intranasal (i.n.) route. Subsequently, at 48 hours post infection, the mice were treated i.p. with either the parental AVP-21D9at 195 μg/mouse, or with AVP-21D9-PlyG at equivalent number of moles with respect to the parental IgG, corresponding to 216 μg/mouse (n=20 per group). Control mice received buffer vehicle prior to infection (n=10).

### NHP ISTAb evaluation against inhalation anthrax

The NHP study was conducted under biosafety level 3 conditions at Tulane University National Primate Research Center. A total of 14 animals were used in this therapeutic evaluation. Aerosol challenge of the animals was performed using virulent *B. anthracis* spores (*Ames* strain). All animals were aerosol exposed to *B. anthracis* after respiratory measurement by whole-body plethysmography was performed. The aerosol system, target dosing, and exposure dose calculations were performed using published methods (*55*). Thereafter, animals were treated approximately 24h after *B. anthracis* exposure. Five animals were treated (IV infusion) with single dose (20 mg/kg) of AVP-21D9-PlyG (Integrated Biotherapeutics (IBT), Lot # 2184-4A-03 ISTAb-BA01) antibody (Group 1) and another five animals were treated (IV infusion) with single dose (20 mg/kg) of AVP-21D9 (IBT, Lot # 2184-3A-03-BA01) antibody (Group 2). Four animals were sham treated with an irrelevant mAb (IgG121) at equivalent dosage as the ISTAb cohorts. Any animal demonstrating a combination of clinical signs associated with the onset and progression of inhalation anthrax were euthanized. This study was conducted in strict adherence to TNPRC’s approved protocol (IACUC P0499).

### Immunogenicity of Anthrax ISTAb in mouse and NHP

#### Mouse studies

Mice were immunized i.p. (200 µl volume) on day 0 with either AVP-21D9 (n=6) at 330 µg/mouse or AVP-21D9-PlyG (n=6) at equivalent number of moles with respect to the IgG. Sera were collected on day 28, and antibody titers against PA toxins were determined by ELISA following our method described above.

#### NHP studies

On days 42, 70, and 99, samples from the NHP PK study were analyzed for anti-PlyG antibodies in the ISTAb antibodies treated group. IgG treated and pre bleed samples were used as controls. Serum titers were determined based on the standard ELISA protocol using HRP-conjugated anti-human IgG for detection as described in the following PK NHP section.

### Pharmacokinetic (PK) analysis in mouse and NHP

#### Mouse studies

Six ICR mice/group were treated i.p. (200 µl volume) with either 330 µg/mouse IgG or equivalent number of moles of derivative ISTAb on day 0. Because IACUC protocols (#161007) only allowed 10% blood loss within 2 weeks, animals were further divided into two groups. One subgroup was bled on days 3 and 6, and the other group was bled on days 14 and 21. On day 28, both groups were terminally bled. Quantitative ELISA was used as described above to determine half-life of the antibodies/ISTAbs.

#### NHP studies

##### I. Dosing

Briefly, four Rhesus macaques were treated (IV infusion) with single dose (20 mg/kg) of AVP-21D9-PlyG antibody (Group 1) and AVP-21D9 (Group 2). Serum samples from day 0, 1, 2, 4, 7, 10, 14, 42, 70, and 99, were used to conduct PK and immunogenicity studies.

##### II. Quantitative ELISA

Quantitative ELISA was used in this study to measure the concentration of antibody in NHP serum samples, as described previously (*56*). Briefly, ELISA plates were coated with 100 ng/well of PA and the concentration of antibody (μg/ml) in NHP serum samples were calculated by comparing absorbance with standard antibody.

##### III. Data analysis

SoftMax Pro 7.1 software was used to analyze the data. Figures were generated by plotting data using software GraphPad Prism 9. Macros PK solver 2.0 was used to calculated T1/2, and C_max_ of AVP-21D9-PlyG and AVP-21D9 control.

## Supporting information

C. difficile Supplemental Methods

## Data Availability

All data produced in the present work are contained in the manuscript

