## Supplementary material for "Engineered antibodies targeted to bacterial surface integrate effector functions with toxin neutralization to provide superior efficacy against bacterial infections": C. difficile Supplemental Methods

+ equally contributed

[1]Integrated Biotherapeutics Inc., Rockville, MD 20850; [2]School of Systems Biology, and the Center for Infectious Disease Research, George Mason University, Manassas, VA 20110; [3]Institute for Bioscience and Biotechnology Research, University of Maryland, Rockville, MD 20850; [4]Institute for Therapeutic Innovation (ITI), University of Florida, Orlando, FL 32827; [5]Tulane University National Primate Research Center, Covington, LA 70433; [6]Department of Veterinary Medicine, University of Maryland, College Park, MD 20742; [7]Center for Aerosol Infection & Transmission Science, Tulane School of Medicine, New Orleans, LA 70112

*Corresponding authors:





Funding: Supported by NIH award AI122666 to RPA, DCN, and RMH; NIH award OD011104 CJR

**Keywords:** Anthrax, Protective antigen (PA), ISTAb, *Bacillus anthracis*, *C. difficile, TcdA, TcdB* AVP-21D9, Toxin Neutralization, Therapeutic

### Supplemental Methods

#### Supplemental Method 1: Identification of *C. difficile* CWTs for ISTAb construction

Bioinformatic screening for endolysins from *C. difficile* phage was carried out and ~25 endolysin or endolysin-like sequences were analyzed using PFAM database to identify catalytic domains (i.e., EADs) and define domain boundaries. Endolysins that lacked known CBD domains or had less than 50 non-catalytic domain amino acids were excluded from further analysis. Eventually, 6 endolysin CDBs (630A, QCD, CD27L, C2, CD119, and CCL2), which ranged in homology from 30% to 90% identity, were selected for further study.

#### Supplemental Method 2: Seroprevalence of *C. difficile* CWT domains in healthy humans

Forty-seven human serum samples from healthy donors (Emergent BioSolutions plasmapheresis program) (*1*) were assessed for pre-existing antibody titers against the *C. difficile* CWTs CCL2 and 630A. As controls, we used the *S. aureus* toxic shock syndrome toxin 1 (TSST-1) as a positive control and the purified glycoprotein (GP) of Ebola virus as negative control. Proteins (100 ng/well) were coated in NUNC ELISA plates overnight at 4°C. Plates were blocked for 1 h at room temperature. Serum samples were diluted at 1:300 in PBS and detected using anti-human IgG-HRP. The $OD_{650nm}$ values were recorded and analyzed using Spectramax 190 plate reader (Molecular Devices) and Softmax 5.4.5 software. $EC_{50}$ values were determined using 4PL plots.

#### Supplemental Method 3: Selection and construction of *C. difficile* IgGs and ISTAbs

We selected well characterized humanized monoclonal antibodies anti-TcdA (6G5) and anti-TcdB (9H10) to generate ISTAb prototypes with the 630A and CCL2 CWTs. The heavy chain of each antibody was fused with the CWTs and co-expressed with the respective light chain to generate TcdA-630A-ISTAb, TcdA-CCL-ISTAb, TcdB-630A-ISTAb, and TcdB-CCL-ISTAb.

#### Supplemental Method 4: ELISA for *C. difficile* IgG/ISTAb binding to toxin/bacteria

The four candidate ISTAbs were tested to confirm they retained the *C. difficile* binding specificity exhibited by the parental CWTs. Eleven *C. difficile* strains with different genotypic and phenotypic backgrounds, along with 12 control strains other than *C. difficile*, were included in the study (Fig.S6C). Binding of the ISTAbs in comparison to the parental mAbs was tested by ELISA using TcdA and TcdB toxins as coating antigen.

#### Supplemental Method 5: Toxicity and toxin neutralization assays

*Toxicity assay*: A cell based toxicity assay for TcdA and TcdB was developed. TcdA and TcdB, once in the cytosol of target cells, inactivate small GTPases such as Rho, Rac, and Cdc42 by glucosylation in the GTP binding site, leading to actin condensation, cell rounding, and death. Inactivation of small GTPases triggers apoptotic pathways through activation of capspase-3 and caspase-9 (*2*). We developed and optimized a cytotoxicity assay for TcdA and TcdB by measuring caspase activity using Caspase-Glo 3/7 (Promega) based on a previously published method with some modification (*3*). Dilutions of toxin are added to Vero cells seeded in 96 wells plates and incubated at 37˚C; 5% $CO_2$ for 24 hours. Caspase-Glo 3/7 buffer (equilibrated to room temperature) is mixed with Caspase Glo 3/7 substrate in a dim light environment and added to the cells at 100 µl/well. Plates are placed in a dark environment for 30 minutes and then read using a BioTek Cytation 5 Image Reader.

*Toxin neutralization assay (TNA)*: Toxin concentrations were determined that result in 90-100% cell lysis based on dose response curve (Fig S6G). To determine neutralizing titer, 25 µl of serial dilutions of either parental IgGs or ISTAbs were incubated with 25 µl of TcdA or TcdB for 15-20 minutes before adding to Vero cells, and the assay was run as above. Toxin neutralization was calculated as % neutralization = (% viable cells treated with IgG or ISTAb and toxin)/(% viable cells without treatment) x 100.
