## Supplemental Tables for "Engineered antibodies targeted to bacterial surface integrate effector functions with toxin neutralization to provide superior efficacy against bacterial infections"

+ equally contributed

[1]Integrated Biotherapeutics Inc., Rockville, MD 20850; [2]School of Systems Biology, and the Center for Infectious Disease Research, George Mason University, Manassas, VA 20110; [3]Institute for Bioscience and Biotechnology Research, University of Maryland, Rockville, MD 20850; [4]Institute for Therapeutic Innovation (ITI), University of Florida, Orlando, FL 32827; [5]Tulane University National Primate Research Center, Covington, LA 70433; [6]Department of Veterinary Medicine, University of Maryland, College Park, MD 20742; [7]Center for Aerosol Infection & Transmission Science, Tulane School of Medicine, New Orleans, LA 70112

*Corresponding authors:





Funding: Supported by NIH award AI122666 to RPA, DCN, and RMH; NIH award OD011104 CJR

**Keywords:** Anthrax, Protective antigen (PA), ISTAb, *Bacillus anthracis*, *C. difficile, TcdA, TcdB* AVP-21D9, Toxin Neutralization, Therapeutic

**Supplemental Table S1:** Melting temperatures of *B. anthracis* ISTAb/IgG measured by DSF

| Antibody | Tm (°C) | Error (°C) | $\Delta T_m = T_m$ ISTAb - $T_m$ IgG |
|---|---|---|---|
| AVP-21D9 | 67.0 | 0.0 | |
| AVP-21D9-PlyB | 66.5 | 0.0 | -0.5 |
| AVP-21D9-PlyG | 67.3 | 0.4 | 0.3 |
| AVP-21D9-PlyL | 65.7 | 0.3 | -1.3 |
| M18 | 68.5 | 0.5 | |
| M18-PlyB | 55.5 | 0.0 | -13.0 |
| M18-PlyG | 68.3 | 0.3 | -0.2 |
| M18-PlyL | 66.8 | 0.3 | -1.7 |
| W2 | 67.7 | 0.3 | |
| W2-PlyB | 65.0 | 0.5 | -2.7 |
| W2-PlyG | 67.5 | 0.0 | -0.2 |
| W2-PlyL | 67.0 | 0.0 | -0.7 |

**Supplemental Table S2:** Binding to FcγRs for *B. anthracis* IgGs

| **AVP-21D9** | | | | | | | |
|---|---|---|---|---|---|---|---|
| Receptor | $K_D$ | $K_{D2}$ | $K_{on}$ | $K_{on2}$ | $K_{off}$ | $K_{off2}$ | $R^2$ |
| FcγR1A | 4.79E-09 | | 9.00E+04 | | 4.30E-04 | | 0.9953 |
| FcγR2A | 2.40E-08 | | 1.70E+04 | | 4.00E-04 | | 0.9952 |
| FcγR2B | 2.30E-08 | | 2.00E+04 | | 4.60E-04 | | 0.9989 |
| FcγRN | 2.70E-09 | <1E-12 | 6.70E+05 | 4.3E4 | 1.80E-03 | <1.0e7 | 0.9971 |

**Supplemental Table S3:** Binding to FcγRs for *B. anthracis* ISTAbs

| **AVP-21D9-PlyG** | | | | | | | |
|---|---|---|---|---|---|---|---|
| Receptor | $K_D$ | $K_{D2}$ | $K_{on}$ | $K_{on2}$ | $K_{off}$ | $K_{off2}$ | $R^2$ |
| FcγR1A | 8.80E-09 | 1.70E-09 | 3.00E+04 | 2.20E+05 | 3.70E-04 | 5.80E-06 | 0.9997 |
| FcγR2A | 3.50E-08 | | 4.50E+04 | | 1.60E-03 | | 0.9959 |
| FcγR2B | 3.50E-08 | 9.10E-08 | 9.10E+03 | 2.80E+05 | 3.20E-04 | 2.60E-02 | 0.9997 |
| FcγRN | 2.00E-09 | <1E-12 | 6.60E+05 | 8.30E+04 | 1.30E-03 | <1.0e7 | 0.9985 |

**Supplemental Table S4:** NHP PK studies (Summary of PK parameters)

| Parameter | Ab dosed (20 mg/kg) | |
|---|---|---|
| | **AVP-21D9** | **AVP-21D9-PlyG** |
| Lambda_z (1/d) | 0.08 | 0.16 |
| T 1/2 (d) | 8.44 | 4.21 |
| $T_{max}$ (d) | 1.00 | 1.00 |
| $C_{max}$ (µg/ml) | 216.62 | 73.88 |
| C0 (µg/ml) | 231.87 | 117.05 |
| AUC 0-t (µg/ml*d) | 2744.93 | 338.88 |
| AUC 0-inf_obs (µg/ml*d) | 2755.59 | 339.00 |
| AUC 0-t/0-inf_obs | 1.00 | 1.00 |
| AUMC 0-inf_obs (µg/ml*d^2) | 27661.76 | 1246.97 |
| MRT 0-inf_obs (d) | 10.03 | 3.68 |
| Vz_obs ((mg/kg)/(µg/ml)) | 0.088 | 0.36 |
| Cl_obs (mg/kg)/(µg/ml)/d | 0.007 | 0.06 |
| Vss-obs (mg/kg)/(µg/ml) | 0.072 | 0.22 |

AUC0-t , AUC from 0 to t h; z, elimination constant; CL, clearance; Vz, apparent volume of distribution during terminal phase; MRT, mean residence time; $T_{max}$, time to maximum concentration of drug in serum.
