## Supplemental figures for "Engineered antibodies targeted to bacterial surface integrate effector functions with toxin neutralization to provide superior efficacy against bacterial infections"

+ equally contributed.

[1]Integrated Biotherapeutics Inc., Rockville, MD 20850; [2]School of Systems Biology, and the Center for Infectious Disease Research, George Mason University, Manassas, VA 20110; [3]Institute for Bioscience and Biotechnology Research, University of Maryland, Rockville, MD 20850; [4]Institute for Therapeutic Innovation (ITI), University of Florida, Orlando, FL 32827; [5]Tulane University National Primate Research Center, Covington, LA 70433; [6]Department of Veterinary Medicine, University of Maryland, College Park, MD 20742; [7]Center for Aerosol Infection & Transmission Science, Tulane School of Medicine, New Orleans, LA 70112

*Corresponding authors:

Funding: Supported by NIH award AI122666 to RPA, DCN, and RMH; NIH award OD011104 CJR

**Keywords:** Anthrax, Protective antigen (PA), ISTAb, *Bacillus anthracis*, *C. difficile, TcdA, TcdB* AVP-21D9, Toxin Neutralization, Therapeutic

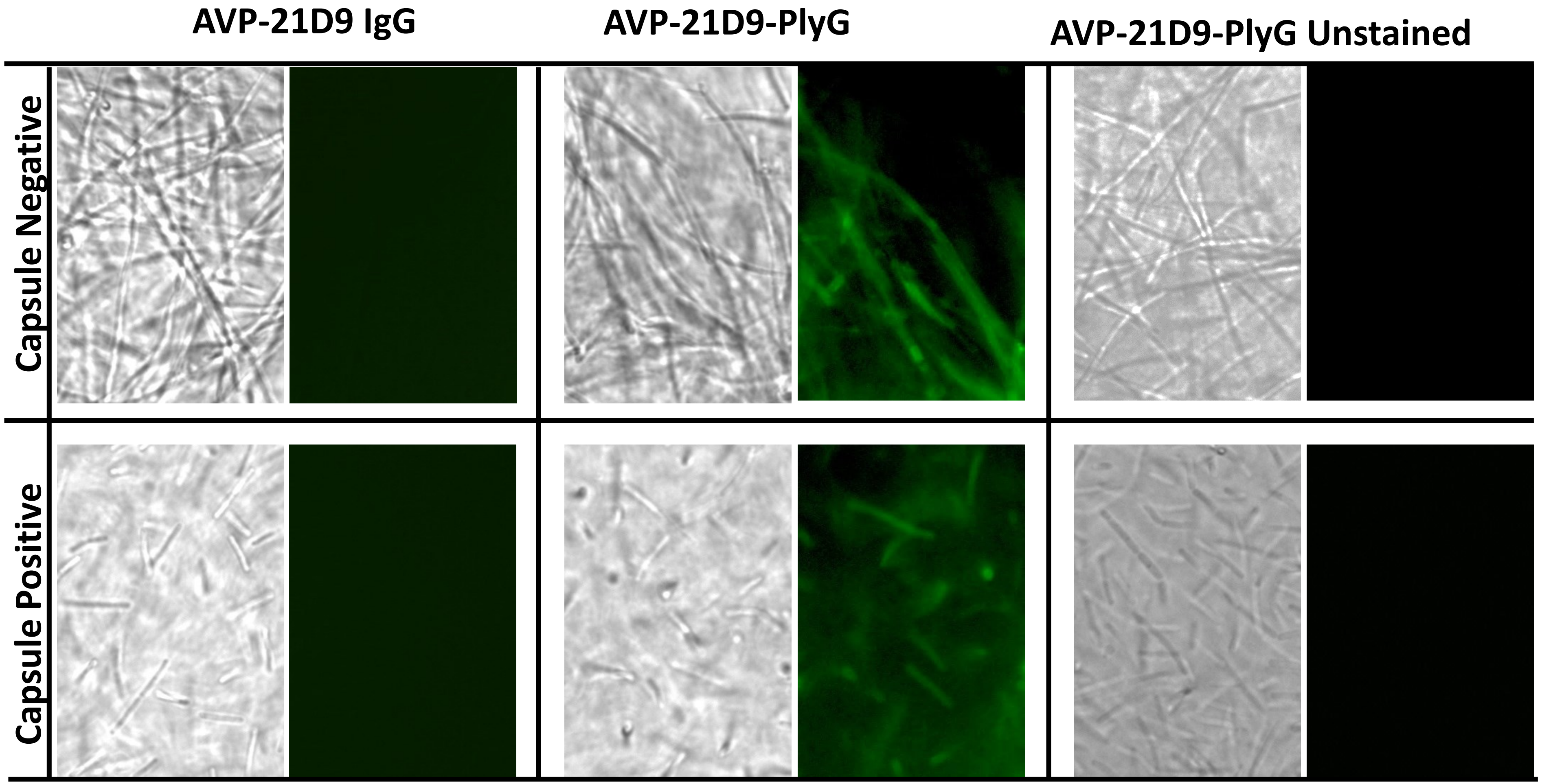


**Fig. S1: ISTAb binding to capsulated bacteria**. Bacteria were grown in either regular media, or in bicarbonate-containing media for capsule formation, and binding of either parental IgG (AVP-21D9) or its derivative ISTAb (AVP-21D9-PlyG) to bacteria was visualized using Alexa 488-conjugated secondary antibody. As a second control condition, bacteria were incubated with the ISTAb without subsequent addition of the secondary antibody (AVP-21D9-PlyG Unstained).

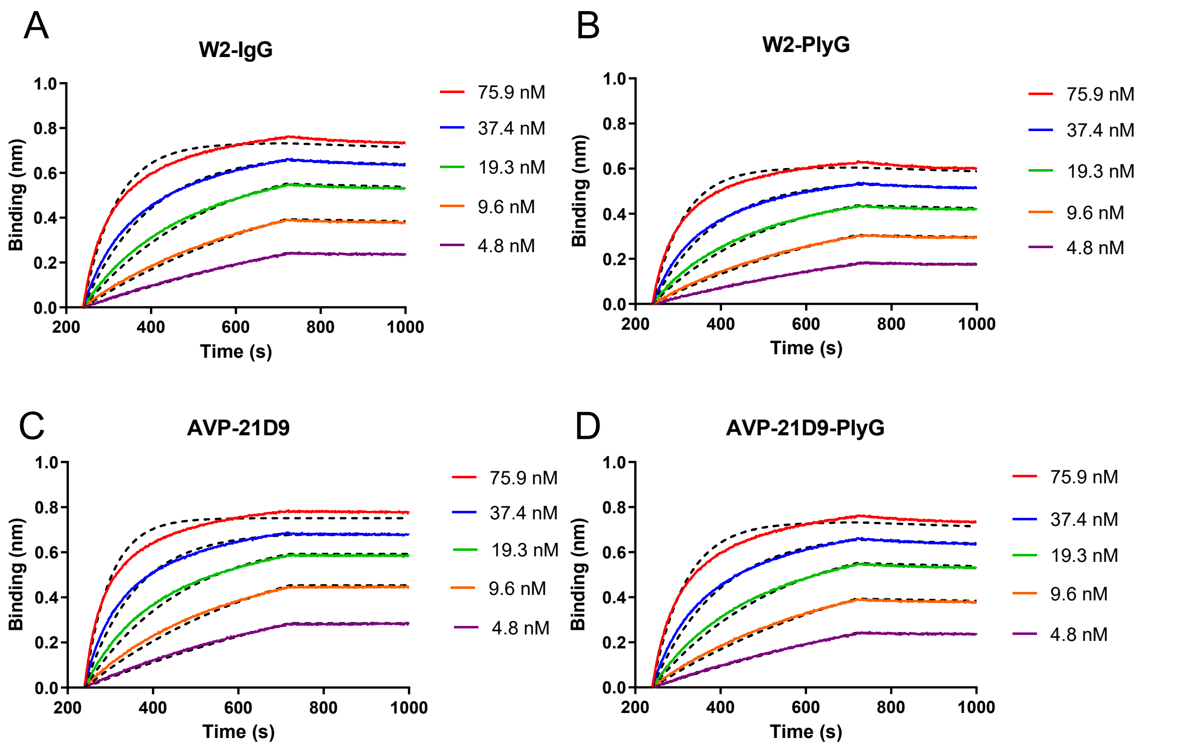


**Fig. S2: Binding kinetics of *B. anthracis* IgGs and ISTAbs to PA.** Binding kinetics and fitting curves are shown for W2 **(A)** and AVP-21D9 (**C**), and their respective ISTAbs **(B), (D).** $K_D$ values are presented in Table 1.

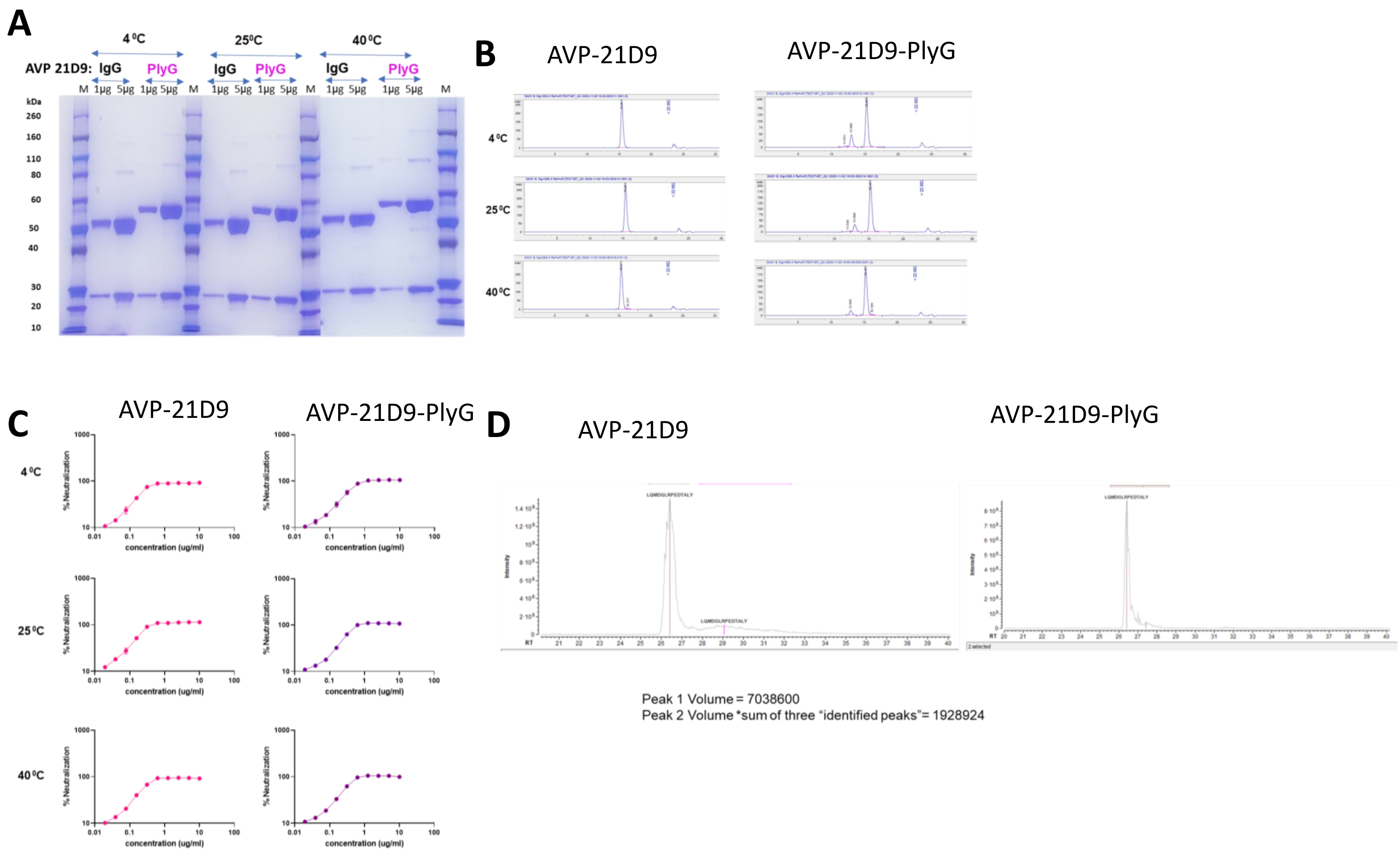


**Fig. S3: Stability of ISTAbs. *B. anthracis*:** Stability of the AVP-21D9 parental IgG versus its derivative PlyG ISTAb, stored for 1 month at the indicated temperatures; **(A)** SDS-PAGE analysis (1 and 5 µg loaded); **(B)** SEC-HPLC analysis; **(C)** TNA activity assay of AVP-21D9 and its derivative PlyG ISTAb after storage for one month at various temperatures. **(D)** LCMS analysis for AVP-21D9 IgG and AVP-21D9-PlyG.

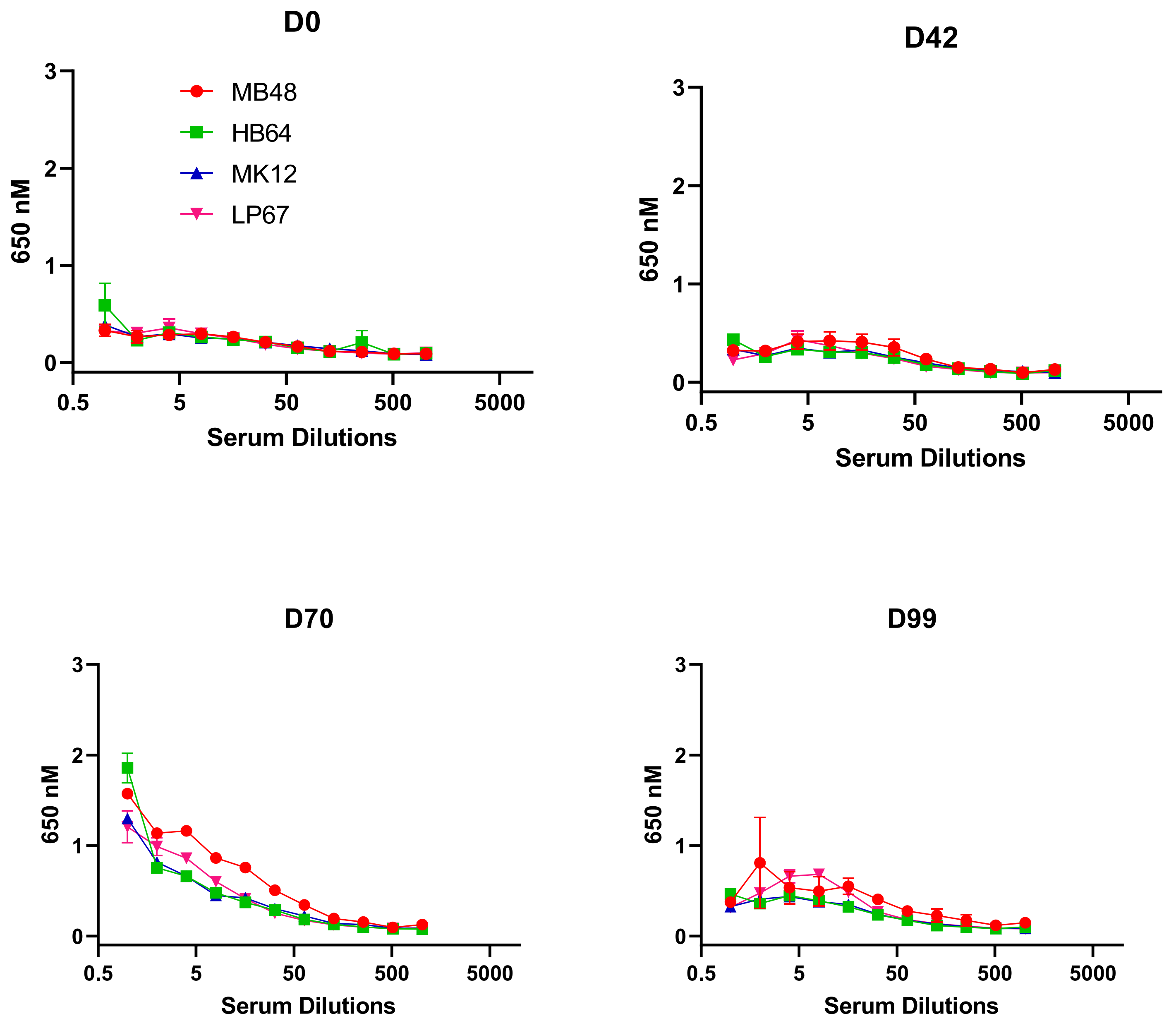


**Fig. S4: NHP immunogenicity studies**. Four NHPs were treated with 20 mg/kg of AVP-21D9-PlyG and serum samples at days D0, D42, D70, and D99 were analyzed for anti-PlyG antibodies. Legends are the same as shown in the D0 across D42, D70 and D99.

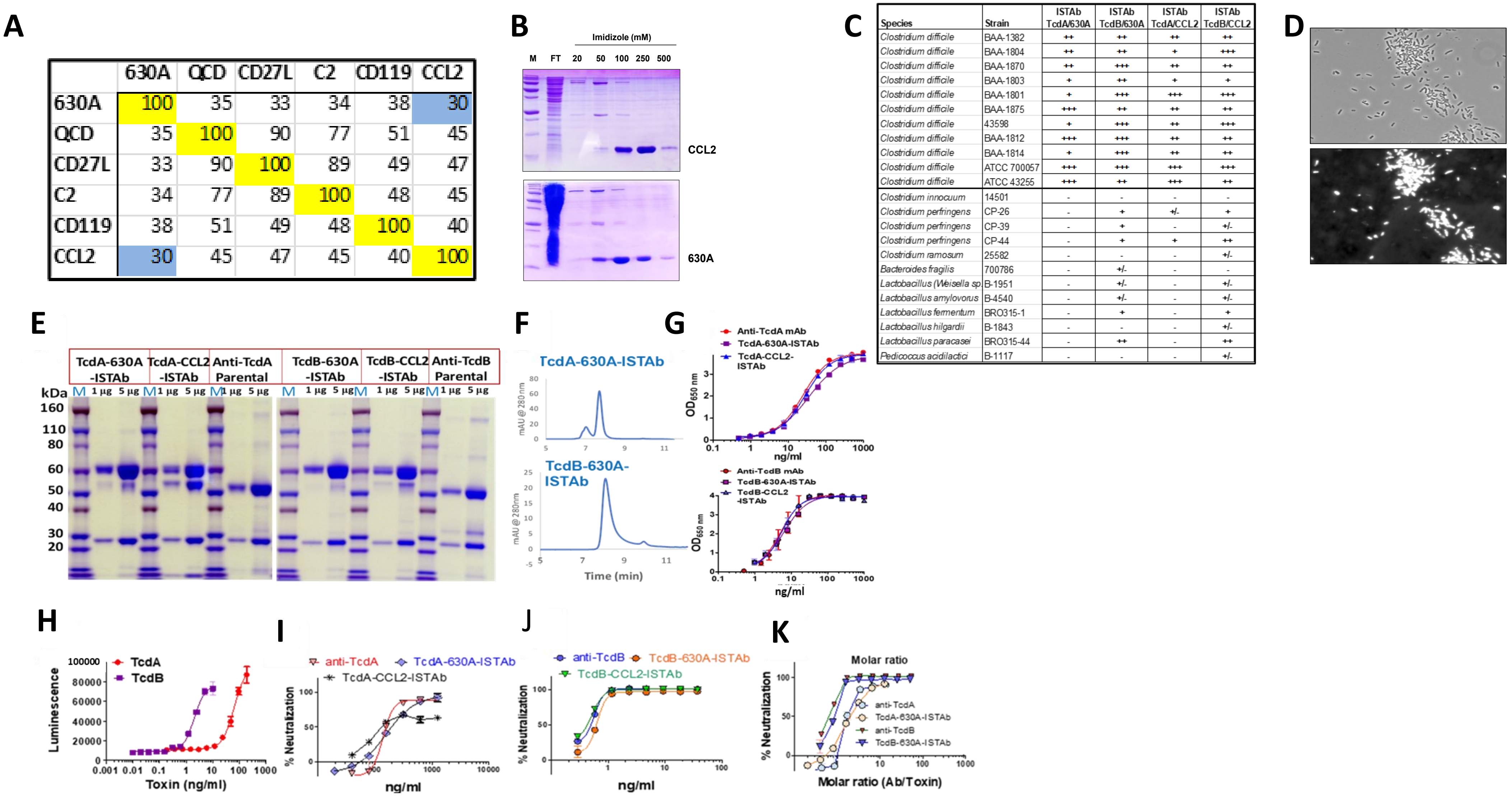


| | 630A | QCD | CD27L | C2 | CD119 | CCL2 |
|---|---|---|---|---|---|---|
| 630A | 100 | 35 | 33 | 34 | 38 | 30 |
| QCD | 35 | 100 | 90 | 77 | 51 | 45 |
| CD27L | 33 | 90 | 100 | 89 | 49 | 47 |
| C2 | 34 | 77 | 89 | 100 | 48 | 45 |
| CD119 | 38 | 51 | 49 | 48 | 100 | 40 |
| CCL2 | 30 | 45 | 47 | 45 | 40 | 100 |

| Species | Strain | ISTAb TcdA/630A | ISTAb TcdB/630A | ISTAb TcdA/CCL2 | ISTAb TcdB/CCL2 |
|---|---|---|---|---|---|
| *Clostridium difficile* | BAA-1382 | ++ | ++ | ++ | ++ |
| *Clostridium difficile* | BAA-1804 | ++ | ++ | + | +++ |
| *Clostridium difficile* | BAA-1870 | ++ | +++ | ++ | ++ |
| *Clostridium difficile* | BAA-1803 | + | ++ | + | + |
| *Clostridium difficile* | BAA-1801 | + | +++ | +++ | +++ |
| *Clostridium difficile* | BAA-1875 | +++ | ++ | ++ | ++ |
| *Clostridium difficile* | 43598 | + | +++ | ++ | +++ |
| *Clostridium difficile* | BAA-1812 | +++ | +++ | ++ | ++ |
| *Clostridium difficile* | BAA-1814 | + | +++ | ++ | ++ |
| *Clostridium difficile* | ATCC 700057 | +++ | +++ | +++ | +++ |
| *Clostridium difficile* | ATCC 43255 | +++ | ++ | +++ | ++ |
| *Clostridium innocuum* | 14501 | - | - | - | - |
| *Clostridium perfringens* | CP-26 | - | + | +/- | + |
| *Clostridium perfringens* | CP-39 | - | + | - | +/- |
| *Clostridium perfringens* | CP-44 | - | + | + | ++ |
| *Clostridium ramosum* | 25582 | - | - | - | +/- |
| *Bacteroides fragilis* | 700786 | - | +/- | - | - |
| *Lactobacillus (Weisella sp.* | B-1951 | - | +/- | - | +/- |
| *Lactobacillus amylovorus* | B-4540 | - | +/- | - | +/- |
| *Lactobacillus fermentum* | BRO315-1 | - | + | - | + |
| *Lactobacillus hilgardii* | B-1843 | - | - | - | +/- |
| *Lactobacillus paracasei* | BRO315-44 | - | ++ | - | ++ |
| *Pedicoccus acidilactici* | B-1117 | - | - | - | +/- |

**Fig. S5:** ***C. difficile* ISTAbs constructions and characterizations. *C. difficile* CWTs:** **(A)** Sequence identity of six *C. difficile* endolysin CWTs. The CWTs for 630A and CCL2 were selected for ISTAb construction due to their binding properties and low homology (30%). **(B)** Purification of CCL2 and 630A CWTs by imidazole elution of Ni-NTA column. **(C)** The ability of *C. difficile* candidate ISTABs to bind target bacteria, closely related species, and anaerobic organisms known to colonize the gastrointestinal tract, was measured by fluorescence microscopy **(D)** Representative binding of the 630A CWT to *C. difficile* BAA-1812. Top: Bright-field image. Bottom: Fluorescent image**. Characterization of ISTAb and IgG. (E)** SDS-PAGE; **(F)** HPLC; and **(G)** ELISA analysis of ISTAbs and parental mAbs. **ISTAb and IgG Toxicity and TNA. (H)** Dose-response of TcdA and TcdB in the caspase assay. Toxin neutralization by ISTAbs and parental mAbs against TcdA **(I)** and TcdB **(J)** respectively. **(K)** Data from **(H)** and **(I)** presented with the molar ratio of antibody/toxin on the X axis**.**

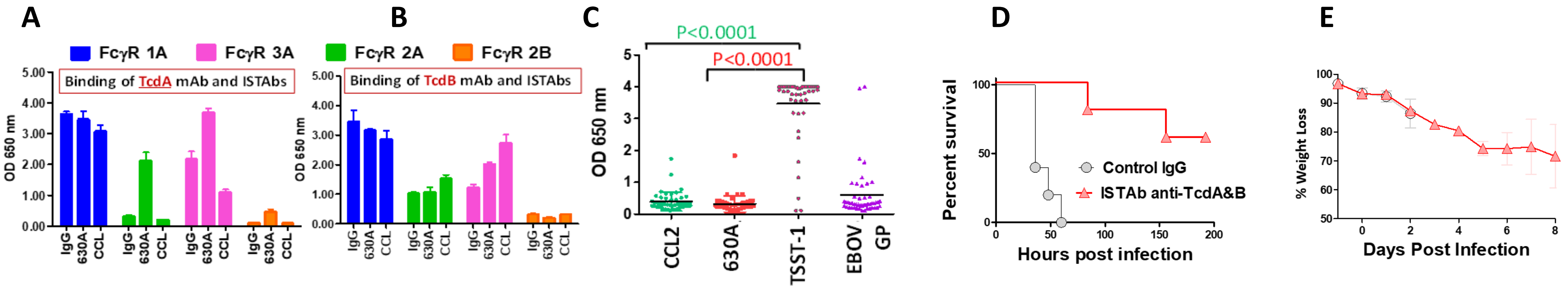


**Fig. S6: *C. difficile* ISTAbs data.** Binding of *C. difficile* ISTAbs to Fcγ receptors. Binding of recombinant Fc receptors to parental IgG, 630A-ISTAb (630A), or CCL2-ISTAb (CCL) for **(A)** TcdA and **(B)** TcdB. **(C)** Serum reactivity to *C. difficile* CWTs. ELISA reactivity of 47 human serum samples to the indicated antigens was measured at 1:200 dilution. Statistical analysis was performed using a t-test, and the calculated p values are indicated. **Hamster efficacy studies with *C. difficile* ISTAb.** Clindamycin (10 mg/mouse) was administered IP one day prior to the challenge, and antibodies (either IgG control, or TcdA and TcdB_ISTAb cocktail) at 1 mg/Hamster were administered for four days starting one day prior to the challenge. Hamsters were monitored for 7 days p.i. **(D)** Survival plot. **(E)** Weight loss in (%).

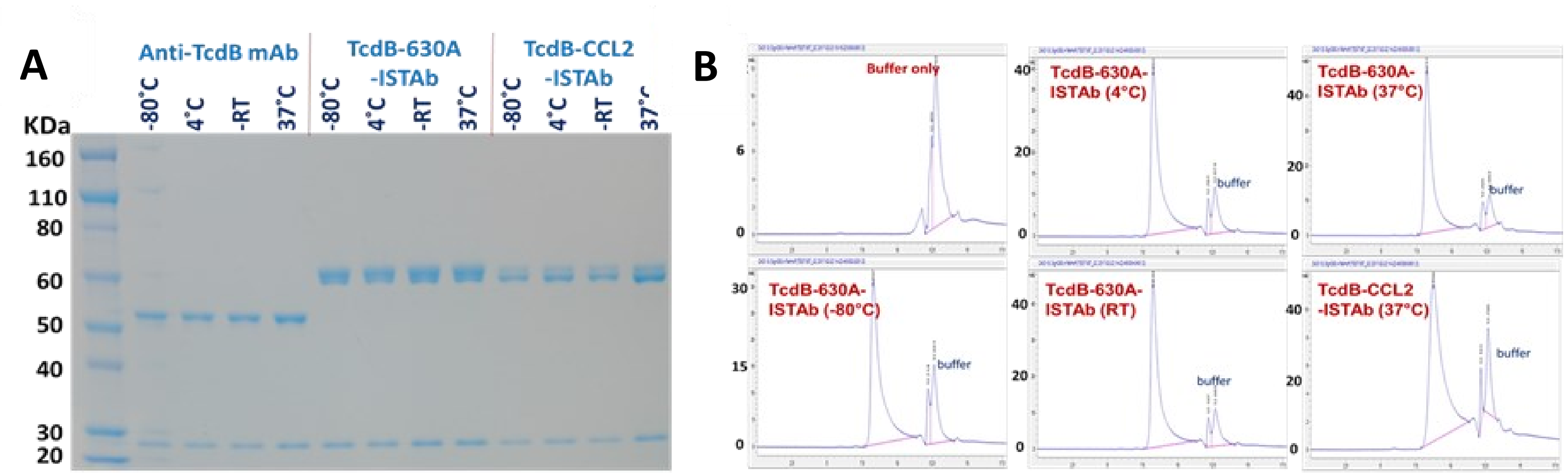


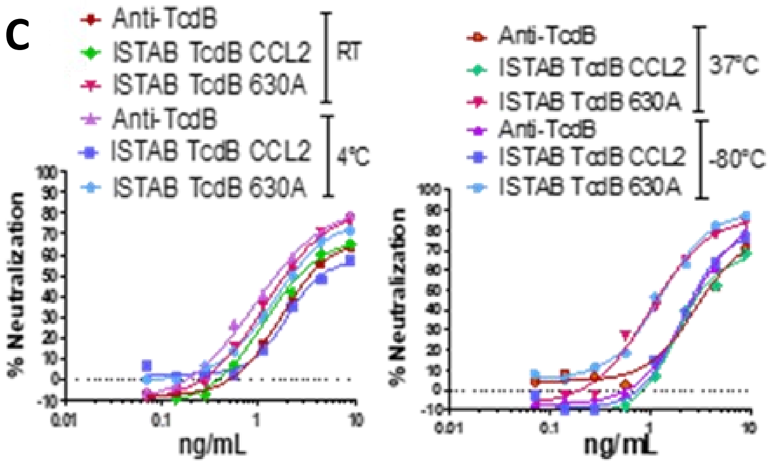


| IC50 (ng/ml) after 1 month Storage | | | |
|---|---|---|---|
| Temperature | Parental | ISTAb-630A | ISTAb-CCL2 |
| -80°C | 2.33 | 1.30 | 2.05 |
| -4°C | 0.93 | 1.47 | 1.99 |
| RT | 1.71 | 1.20 | 1.16 |
| -37°C | 2.85 | 0.99 | 1.84 |

**Fig. S7: Stability of ISTAbs. *C. difficile*:** Stability of the ISTAbs and parental TcdB IgG stored for 1 month at the indicated temperatures; **(A)** SDS-PAGE analysis; **(B)** SEC-HPLC analysis; **(C)** TNA activity of the indicated ISTAbs and IgG after storage for one month at various temperatures

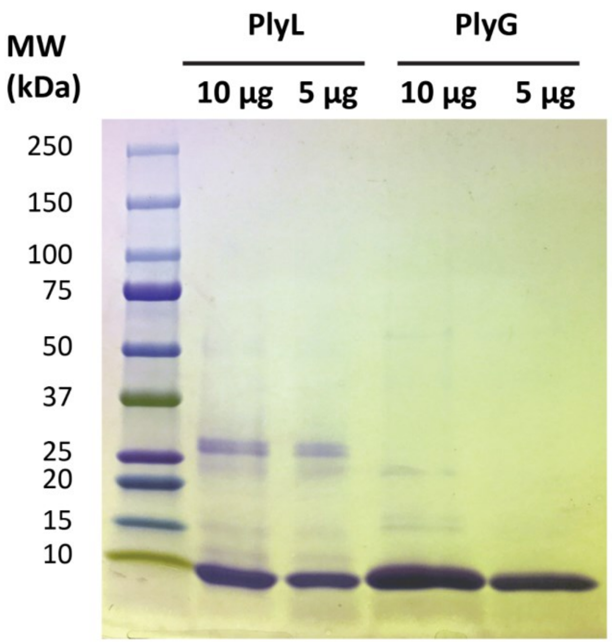


**Fig. S8 Purification of *B. anthracis* CWTs**. The PlyL and PlyG CWTs were expressed in high yields, whereas those of PlyTsamsa and PlyB had very low yields, and PlyAP50 had folding/expression issues.
